# A Clinical Neuroimaging Platform for Rapid, Automated Lesion Detection and Personalized Post-Stroke Outcome Prediction

**DOI:** 10.1101/2025.05.09.25327310

**Authors:** Michal Brzus, Joseph Griffis, Cavan J. Riley, Joel Bruss, Carrie Shea, Hans J. Johnson, Aaron D. Boes

## Abstract

Predicting long-term functional outcomes for individuals with stroke is a significant challenge. Solving this challenge will open new opportunities for improving stroke management by informing acute interventions and guiding personalized rehabilitation strategies. The location of the stroke is a key predictor of outcomes, yet no clinically deployed tools incorporate lesion location information for outcome prognostication. This study responds to this critical need by introducing a fully automated, three-stage neuroimaging processing and machine learning pipeline that predicts personalized outcomes from clinical imaging in adult ischemic stroke patients. In the first stage, our system automatically processes raw DICOM inputs, registers the brain to a standard template, and uses deep learning models to segment the stroke lesion. In the second stage, lesion location and automatically derived network features are input into statistical models trained to predict long-term impairments from a large independent cohort of lesion patients. In the third stage, a structured PDF report is generated using a large language model that describes the stroke’s location, the arterial distribution, and personalized prognostic information. We demonstrate the viability of this approach in a proof-of-concept application predicting select cognitive outcomes in a stroke cohort. Brain-behavior models were pre-trained to predict chronic impairment on 28 different cognitive outcomes in a large cohort of patients with focal brain lesions (N=604). The automated pipeline used these models to predict outcomes from clinically acquired MRIs in an independent ischemic stroke cohort (N=153). Starting from raw clinical DICOM images, we show that our pipeline can generate outcome predictions for individual patients in less than 3 minutes with 96% concordance relative to methods requiring manual processing. We also show that prediction accuracy is enhanced using models that incorporate lesion location, lesion-associated network information, and demographics. Our results provide a strong proof-of-concept and lay the groundwork for developing imaging-based clinical tools for stroke outcome prognostication.

## Introduction

Stroke is a leading cause of long-term disability, with a global incidence of around 12 million new cases per year ^1^. Patients with a stroke typically present with multiple deficits, which may include core cognitive functions like one’s ability to speak or understand language ^2^. While there is often recovery within the first few months, it is common for some deficits to persist and contribute to long-term disabilities. In contrast, other stroke symptoms may not be apparent on initial presentation but develop in the days and weeks after a stroke, such as central post-stroke pain or post-stroke depression. Given the temporal unfolding of stroke sequelae and the high variability in recovery from early deficits, the days and weeks following a stroke can be filled with anxiety and uncertainty for patients and their loved ones.

Clinicians are currently limited in their ability to predict long-term post-stroke outcomes accurately. Research shows that individual clinician prognostication is highly variable and often inaccurate, with significant differences not attributable to experience or subspecialty expertise ^3^. Clinical tools like the NIH Stroke Scale provide some prognostic utility ^4^, but the outcomes predicted are not personalized and lack granularity for the types of deficits that are likely to contribute to disability ^5,6^. More recently, prediction algorithms have been developed for post-stroke motor recovery ^7,8^, but these are limited in that they require steps that are not part of routine clinical practice, and there is no automated way to incorporate lesion anatomy into the prognostication. These algorithms only address motor outcomes and do not provide a generalizable framework that can be applied to cognition and other outcomes that significantly influence quality of life and disability. Despite widespread recognition of the need for evidence-based clinical prognostication tools ^9^, no tools that can aid clinicians in making personalized, post-stroke predictions of functional outcomes currently exist.

Developing a clinically viable tool for automated outcome prognostication could help patients and clinicians navigate post-stroke life more effectively. An automated tool to inform outcome prognostication could help guide management decisions in the inpatient setting, inform rehabilitation interventions, and help plan more effectively for the level of services that may be required. Accurate prognostic information could also inform personalized anticipatory guidance, such as education and increased surveillance of post-stroke pain or mood symptoms for individuals at higher risk for these symptoms. Finally, providing outcome information to patients shortly after having a stroke could motivate earlier initiation of rehabilitation when patients may be most likely to respond.

It is critical to account for the stroke anatomy to enable accurate prognostication. For instance, a lesion that involves the corticospinal tract is strongly associated with poorer recovery of motor function ^10,11^. In contrast, lesions that spare this tract tend to have a much better motor prognosis. The same principle applies to brain structures and networks serving other functions. Work by our group and by others has identified many critical anatomical regions and distributed networks associated with different types of impairments ^2,12–25^. Importantly, statistical models can be trained to leverage lesion location, lesion-associated networks, and other non-lesion factors (e.g. demographic information) to generate personalized outcome predictions ^17,26–28^.

Despite these advances in lesion research for prognostication, to our knowledge, no tools have been developed in a way that would be suitable for clinical implementation. Existing approaches require specialized image processing steps and manual interventions that make them unsuitable for clinical use. As a result, the well-established corpus of knowledge about lesion-behavior relationships has not been successfully translated into practical clinical applications.

To address this gap, we present a framework for an automated clinical tool that can leverage raw clinical imaging to generate personalized and anatomically informed outcome predictions in acute ischemic stroke, building upon our previous work ^18,27,29^. Starting with raw MRI data routinely acquired for the diagnosis of stroke, our platform automatically identifies and segments the stroke lesion and uses the lesion location, derived network features, and patient demographic information as inputs to pre-trained machine learning models that generate predictions about long-term neuropsychological outcomes. A personalized report is automatically generated for each run that describes the lesion location and its arterial distribution, and that also visualizes the stroke and lesion segmentation for clinician verification.

As a proof of concept, we trained brain-behavior models to predict impairment status on an array of neuropsychological outcome measures spanning different cognitive domains in a large cohort of adult patients with brain lesions (N=604), embedded them into the platform, and applied the complete end-to-end prototype tool to clinical imaging data from an independent test cohort of patients with ischemic stroke (N=153), obtaining personalized outcome predictions for each patient in the independent cohort. We demonstrate robust performance of the processing pipeline across diverse clinical scanners and acquisition sequences. We also show that our anatomically informed modeling approach can achieve meaningful and statistically significant outcome prediction performance when applied to independent clinical imaging datasets, and that incorporating additional information about affected networks and patient demographics can improve prediction performance beyond that afforded by lesion location alone. Altogether, this work provides strong proof of concept for our approach and represents an important step toward implementing tools for automated, imaging-guided personalized outcome prediction in acute stroke care.

## Materials and Methods

The methods are described in three parts: **Part I** describes image processing and lesion segmentation, **Part II** describes a cognitive outcome prediction module, and **Part III** outlines the report generation system, which combines large language model (LLM) technology to create clinically relevant summaries describing lesion location, arterial distributions, and predicted outcomes. **Figure 1** shows the major steps of the processing flow across these three major components. **Supplemental Figure 1** shows a more detailed visualization of MRI processing for those steps.

**Figure 1.**
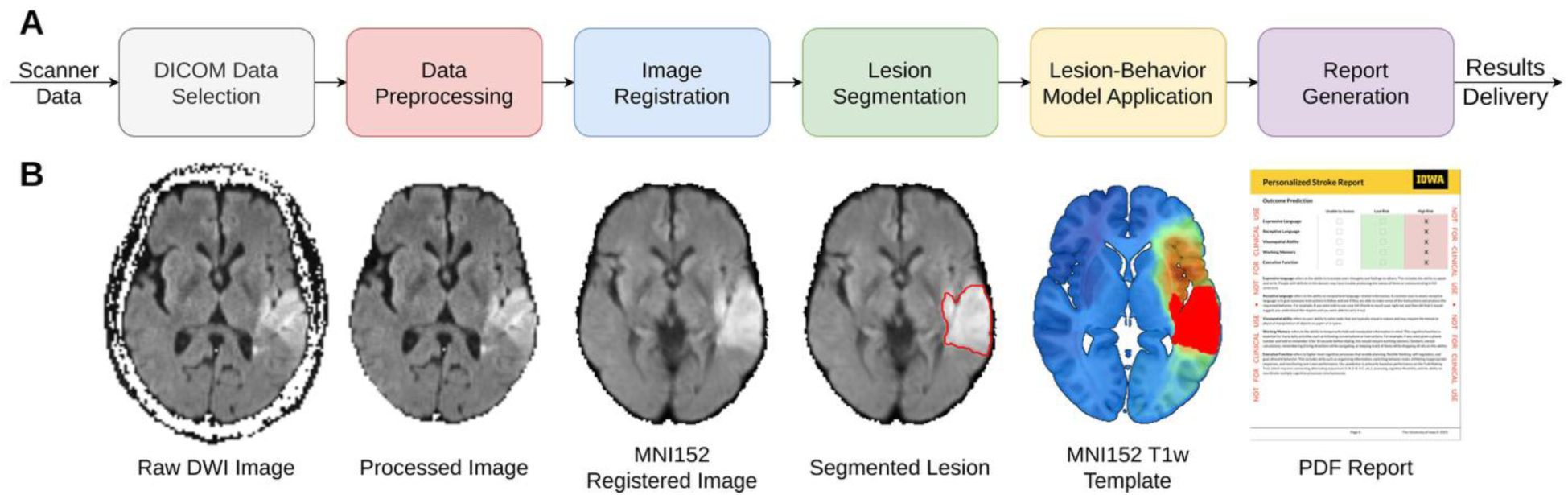
Overview of the personalized stroke outcome prediction pipeline. **A.** Diagram illustrating the end-to-end data flow, from raw scanner data to the final prediction report, highlighting key system components. **B.** Visualization of the diffusion weight imaging sequence at various processing stages, including the unprocessed image, processed image, MNI152- registered image, segmented lesion, MNI152 T1w template with lesion-symptom map and binary lesion mask, and the final PDF report. Detailed descriptions of each step are provided in the text and **Supplementary Material.**

### Part I. Image Processing and Lesion Segmentation Module

The individual pipeline components included in Part 1 involve selecting and processing raw clinical MRI data, preprocessing, template registration, and lesion segmentation. The minimum system requirements include one structural image—either T1-weighted (T1w), T2-weighted (T2w), or Fluid-Attenuated Inverse Recovery (FLAIR)—along with Diffusion Weighted Image (DWI) Trace and Apparent Diffusion Coefficient (ADC) images, which are imaging sequences routinely acquired in the early management of stroke. The development and validation of engineering solutions within this section utilized multiple datasets, with detailed methodology and experimental results provided in the **Supplementary Material**, some of which have been published previously. Here, we evaluate the complete system on an independent, out-of-sample cohort of patients with acute clinical imaging (N=160) and chronic outcome measures (N=153). This test dataset is described in **Part 2,** and the performance of the system on the test dataset is presented in the results section.

**DICOM Data Selection.** Pinpointing the specific subset of DICOM objects acquired during a scanning session is a fundamental, yet frequently neglected challenge in integrating automated analysis tools into clinical settings. To ensure the downstream success of the various algorithms in the pipeline, different image modalities (structural/anatomical sequence, DWI, and ADC) must be reliably identified in a fully automated way. For this purpose, we utilized our open-source *dcm_classifie*r package. This extensible classification framework extracts key features from well-characterized DICOM header fields to identify image modality and acquisition plane with over 99% accuracy ^30^.

For the current pipeline, we extended the tool’s capabilities with post-processing functionality that encompasses several critical steps: (1) automatic selection and labeling of individual sequences including T1w, T2w, FLAIR, DWI, and ADC; (2) prioritization of axial and isotropic acquisition planes, which are standard in stroke imaging; (3) verification of sufficient image extent for complete brain coverage; (4) computation of standardized ADC images when sufficient diffusion data is available; and (5) conversion of the images into 3D volumes in NIFTI or NRRD format using ITK ^31^.

This fully automated procedure ensures robust input data selection in a clinical environment. Regardless of manufacturer or model, it is generalizable to any DICOM-conforming MRI scanner, making it highly scalable across different clinical settings.

**Data Preprocessing.** Following image selection, the preprocessing pipeline transforms raw imaging volumes into analysis-ready data through several integrated steps.

First, we generate brain masks for all imaging modalities using a custom-trained 3D ResUNet model ^32^. These masks enable skull-stripping and define regions of interest for subsequent analyses. We trained brain mask models using a three-fold approach: initial model development with 3831 images, hyperparameter optimization with an independent validation set (N=496 images), and final performance assessment on a separate test set (N=496 images) (**Supplementary Table 1**). This model achieved an average Dice coefficient of 0.98 across modalities (**Supplementary Table 2**). Detailed descriptions of datasets and evaluation protocols are provided in the **Supplementary Material**. For quality assurance, we employ the MIQA tool ^33^ to systematically evaluate each image for brain coverage, artifacts, and motion corruption, generating objective quality scores. We then implement a cross-validation framework that compares brain masks across modalities, identifying and excluding outliers through statistical analysis to optimize the anatomical accuracy of the brain masks.

We select the optimal input data by identifying the highest-quality image for each modality (T1w, T2w, FLAIR, ADC, and DWI) that passes the brain mask validation. The highest-scoring anatomical image is selected as the target for subsequent processing, with minimum requirements of one anatomical sequence, DWI, and ADC to proceed.

Finally, we generate a normalized, high-resolution (1mm isotropic) T1w volume from the target image using SynthSR ^34^. This synthesized volume provides standardized contrast properties, enhancing registration reliability and mitigating variability in clinical acquisitions.

**Template Brain Registration.** Registration to the MNI-152 standard brain template enables the application of brain atlases for anatomical description and extraction of prognostic information from statistical models. While stroke lesions can potentially affect registration accuracy, our approach leverages the high-quality synthesized T1w images from the preprocessing step, effectively normalizing pathological changes and improving registration outcomes ^34^. The registration process begins with co-registering the highest quality images from each modality to the synthetic T1w derivative (**Supplementary** Figure 2). We then create a transformation between this synthesized T1w image and the MNI-152 T1w template using a combination of rigid, affine, and non-linear transformations implemented through ITK ^31^ and ANTs ^35,36^, with brain masks defining the registration sampling region (**Supplementary** Figure 3). This single transformation is then applied to all co-registered images, efficiently bringing them into MNI-152 space and resampling them to template dimensions. We performed validation testing on 2,987 individual images as described in the **Supplementary Material**, achieving a 99.7% registration success rate. Notably, the approach using the synthesized T1w image outperformed a conventional approach using the original clinical images **(Supplementary Material**), demonstrating a clear benefit to the use of synthesized T1w images for registration and thus supporting the findings in ^34^.

**Lesion Segmentation.** Accurately segmenting acute ischemic stroke lesions is integral to our pipeline, as outcome prediction models are contingent upon precise lesion identification. The current study builds upon earlier versions of our deep-learning lesion segmentation method ^37^. We trained deep learning models for semantic segmentation on 700 stroke MRI scans, consisting of both clinical scans obtained at the University of Iowa and the ISLES 2022 dataset ^38^, with DWI and ADC sequences used for lesion segmentation. Early models were also trained using additional structural sequences (FLAIR, T1w, or T2w), but analysis did not show significant improvements (**Supplementary Table 3**). Once segmented, the acute infarct is represented as a binary 3D volume in MNI-152 template space (**Figure 2**).

**Figure 2.**
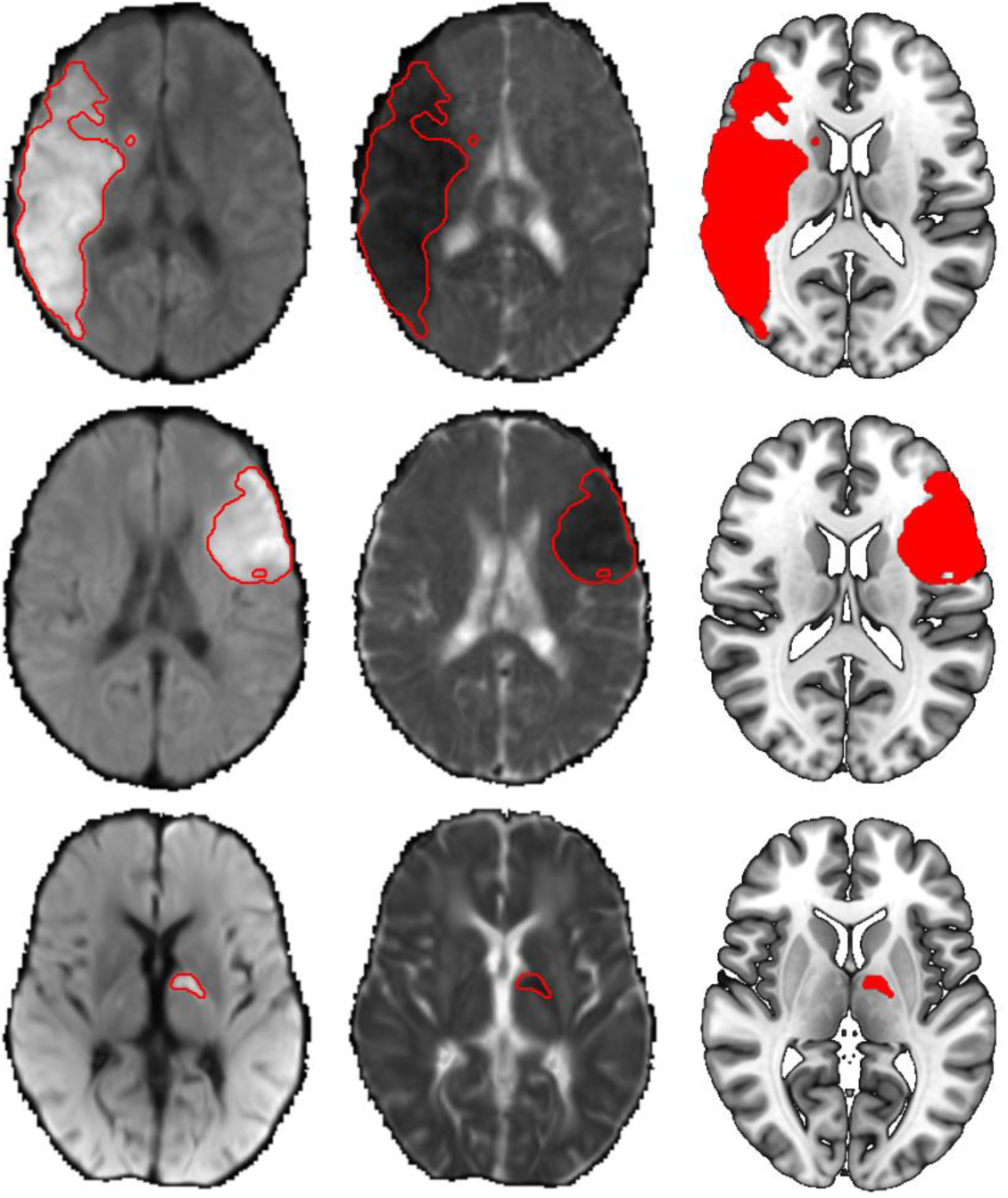
Examples of automated lesion segmentations across three different cases. Axial slices of diffusion weighted imaging (left column) and apparent diffusion coefficient (middle column) images in MNI152 space are input to a Deep Learning model to produce binary lesion masks, outlined in red. The corresponding MNI152 T1w template images (right column) demonstrate the segmented lesions.

**Evaluation of Pipeline Performance and Segmentation.** To evaluate the image processing and lesion segmentation module, we tested its performance on 160 MRIs acquired for the diagnosis and management of stroke within 7 days of the infarct. These scans are independent of the data used to train the lesion segmentation deep learning models. Measures of robustness, accuracy and runtime were all considered. To help evaluate performance, we implemented a custom pipeline profiler class that monitors execution, time, memory, and resource usage in a way that is logged for tracking. A subset of the MRIs in the test cohort was manually traced for comparison to automated lesion segmentation (N=57).

### Part II. Lesion-Informed Cognitive Outcome Prediction

Our overarching goal is to utilize the previously described lesion segmentations to generate individualized predictions of post-stroke functional outcomes. To demonstrate the feasibility of this goal, we used a training dataset to develop statistical models to predict impairment on neuropsychological outcome measures based on lesion location. We applied these models to automatically segmented lesions from an independent test dataset to evaluate prediction performance. Lesion location was the primary input to the models; we also assessed whether performance could be improved by incorporating lesion-derived network features and demographic information (i.e., age and education). The strategy for model training and testing is summarized in **Figure 3**. Inclusion criteria for both training and test cohorts included cognitive outcome data obtained for at least one of 28 cognitive outcomes (**Supplementary Material**), which were selected based on their representation in both the training and test datasets (**Supplementary Table 7**). We included cognitive assessments performed 3 months or later after the lesion onset whenever possible (98.7% of training dataset patients). For the training dataset, patient consent was obtained in accordance with the Declaration of Helsinki. The test dataset was a retrospective sample of convenience pulled from electronic medical records. The need for consent was waived in accordance with the University of Iowa Institutional Review Board policies of using de-identified clinical datasets. The study design and procedures were approved before data collection.

**Figure 3.**
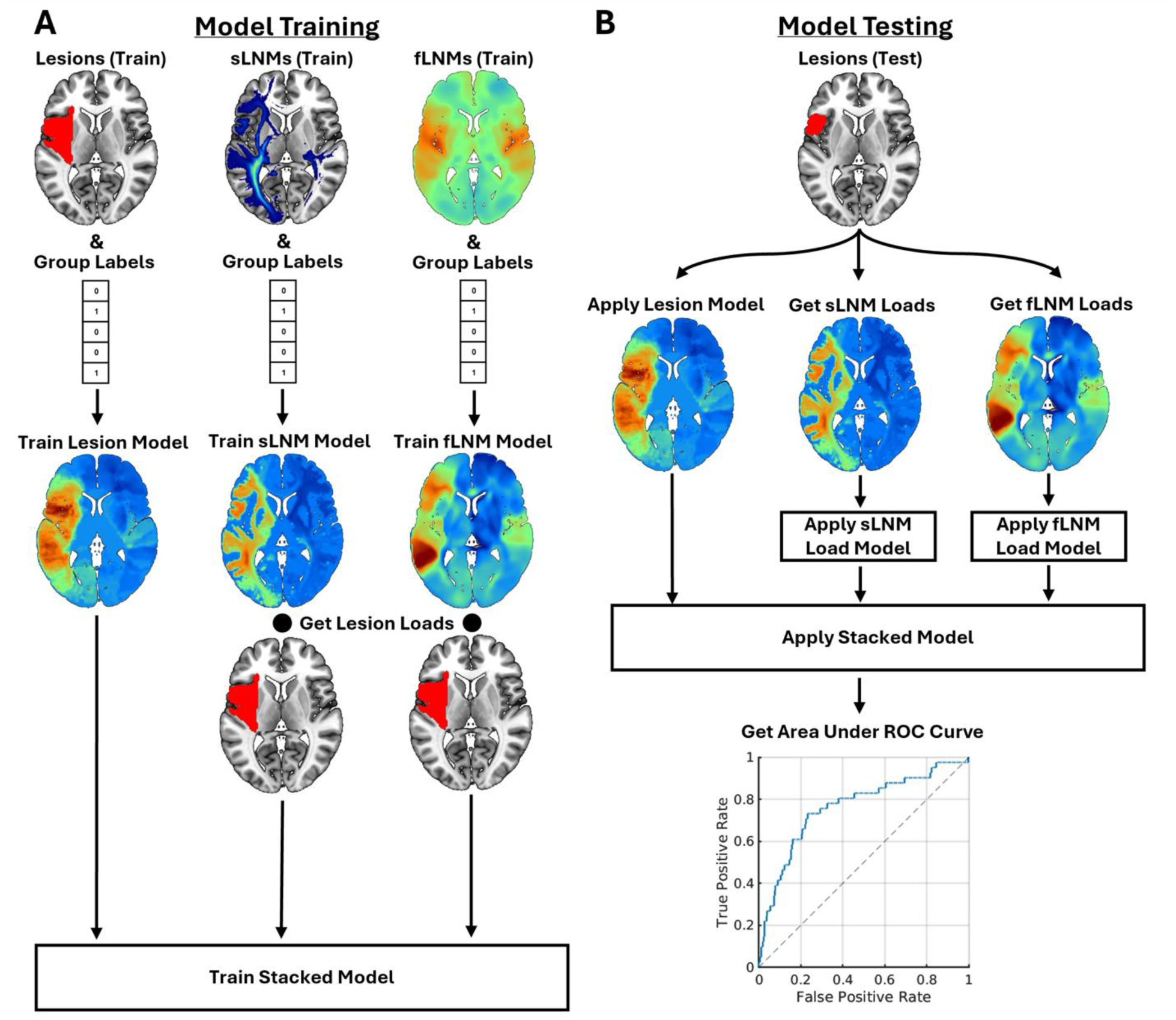
Cognitive outcome prediction modeling methods. **A.** Model training procedure. Partial least squares classification models were trained to classify patients as either ”impaired” or ”unimpaired” using lesion location and derived structural lesion-network map (sLNM) or functional lesion-network map (fLNM) features. A separate model was trained for each feature. For network models, ”lesion load” features were computed as the dot product between the vectorized patient lesion segmentations and vectorized model coefficient maps. Finally, stacked ridge-regularized logistic regression models were trained using the predicted group labels from the lesion models and lesion loads on the sLNM and fLNM models as predictor features, combining information about lesion location and lesion-derived networks. **B.** Model testing procedure. Models were evaluated by applying the trained models to lesion masks and ”lesion load” features (computed on the coefficient maps from the models generated using the training dataset) from the test dataset, obtaining predicted classification scores and predicted group labels for each patient in the test dataset. The predicted classification scores were then used along with the true labels to construct a Receiver Operating Characteristic (ROC) curve, and model performance was evaluated by computing the area under the ROC curve.

**Training Dataset for Cognitive Outcome Prediction.** We identified 604 patients from the Iowa Lesion Registry that met our criteria for inclusion in the training dataset. Patients were included if they had focal, acquired brain lesions and research-quality structural neuroimaging. Patients were excluded from participation in the Registry if they had pre-existing intellectual disability or pre-existing neurological or psychiatric disorders. As mentioned, we identified 28 different cognitive assessments based on their representation in the training and testing datasets and trained classification models to predict impairment status for each of these 28 outcomes.

Training dataset details are provided along with the complete list of cognitive outcome measures and detailed method descriptions in the **Supplementary Material**. Lesion overlap maps are shown in **Supplementary** Figure 4A.

**Testing Dataset for Cognitive Outcome Predictions**. To assess the performance of our proposed system for predicting cognitive outcomes following ischemic stroke, we compiled an independent test dataset of 153 patients with relevant neuroimaging and neuropsychological outcome data. Patients were selected from a retrospective review of clinical records at the Benton Neuropsychology Clinic (University of Iowa Healthcare System), with a confirmed ischemic stroke diagnosis between 2002 – 2023. Inclusion required: (i) clinical MRI data, acquired within one week of stroke onset, including DWI and at least one structural sequence (T1-weighted, T2-weighted, or FLAIR); and (ii) chronic-stage cognitive outcomes, assessed post-stroke via clinical neuropsychology assessment. This cohort is a subset of the 160 used to evaluate the processing and lesion segmentation pipeline. Seven individuals did not have eligible cognitive outcome data and could only be included in the imaging analysis. The imaging data represent substantial heterogeneity, derived from 17 unique scanner models across four manufacturers (Siemens, Philips, GE Medical Systems, and Olea Medical), utilizing 1.5T and 3T field strengths. This diversity of scanners and magnet strength mirrors real-world clinical variability to help evaluate the generalizability of our out-of-sample test results.

Additional details of the test dataset are provided in the **Supplementary Material**. Lesion overlap maps are shown in **Supplementary** Figure 4B.

**Statistical Analyses** We trained multivariate partial least squares (PLS) classification models to classify patients as “impaired” or “unimpaired” in terms of their performance on each of the 28 cognitive outcome measures based on their lesion location, as outlined in **Figure 3** and described in the **Supplementary Material**. Including all 28 cognitive outcomes allowed us to evaluate classification performance across a diverse range of measures, enabled the identification of well-predicted outcome measures that could be prioritized for further development, and enabled statistical comparisons across outcome measures among models with different predictor features in the independent test dataset. Impaired performance was defined using assessment-specific published impairment thresholds when possible; otherwise, when these were unavailable, as a population-normed *z-*score of less than -1.5. For the subset of measures corresponding to clinician ratings of impairment, ratings of 3 (impaired) were considered “impaired”, while ratings of 2 (borderline) and 1 (unimpaired) were considered “unimpaired”. The number of impaired patients in the training and test sets are reported for each measure in **Supplementary Table 7.**

We also used the lesion location information to infer the connectivity of lesioned tissue using normative ‘connectome’ datasets, as performed previously ^18,27,39,40^, and further described in the **Supplementary Material**. We refer to the lesion-derived structural and functional networks as structural lesion network maps (sLNMs) and functional lesion network maps (fLNMs).

Multivariate PLS classification models were trained on lesion-derived network maps using the same approach as lesion location models, producing brain-wide coefficient maps reflecting network topographies associated with impairment. Because patient-specific lesion-network maps are not currently generated within the pipeline, we performed a second modeling step that involved calculating lesion loads as the dot product between each vectorized lesion and each vectorized network model coefficient map, summarizing how each patient’s lesion relates to the predictive structural and functional network topographies. Finally, we combined lesion and network features in “stacked” logistic ridge regression models that included the predicted class labels from the lesion location models and the lesion load features for the sLNM and fLNM models as predictors. We also evaluated the impact of adding age and education information to the stacked models. For these analyses that included demographics, 36 training dataset patients and 10 test dataset patients were excluded due to missing education data.

All models were trained in the training dataset and applied to the independent test dataset. Cross-validation was also performed in the training dataset, as described in the **Supplementary Material.** Performance was measured in terms of the area under the ROC curve (AUC) in the independent test dataset following best-practice recommendations for evaluating classifier performance in neuroimaging studies with imbalanced classes ^41^. We used permutation testing (1,000 permutation iterations) to evaluate the statistical significance of the test set AUC obtained for each outcome measure. Finally, we compared overall performance (i.e., test dataset AUCs across all 28 outcome measures) between the lesion-only models and stacked models using two-tailed Wilcoxon signed rank tests. All modeling was performed using the Iowa Brain-Behavior Modeling Toolkit ^27^. The approaches used for model training and testing are described in detail in the **Supplemental Material**.

### Part III. Report generation

Reports are generated as PDF files using ReportLab and Meta’s LLaMA 3.3 (70B) for narrative synthesis ^42^, combining templated text with imaging-derived insights (e.g., lesion volume, location). The reports synthesize neuroimaging analyses into documentation that could be utilized by clinicians. Clinician-focused sections include an overview, radiological analysis, neuroanatomical analysis, and outcome prediction. The radiological analysis provides visualizations of DWI and ADC sequences with lesion segmentation overlays, alongside 3D renderings in MNI space for spatial context. Visualizations utilized ITK and VTK libraries. The neuroanatomical analysis quantifies lesion overlap with cortical, subcortical, cerebellar, vascular, and white-matter tracts atlases ^43–53^, presenting volumetric intersections across 154 anatomical and 30 arterial territories, 91 white-matter tracts, and 74 thalamic nuclei in hierarchical tables (hemisphere, lobe, subregion). Outcome predictions, personalized for each patient, are displayed as binary (low or high) risk categories for different cognitive domains, enabling rapid assessment of functional impairment risks.

The final two report pages are tailored for patients and families, translating complex medical findings into accessible, non-technical language. These sections cover the stroke event, expected outcomes, risks, and actionable steps, using relatable analogies and avoiding medical jargon. Readability is optimized for a 6th–7th grade level (SMOG metric: mean 6.6, max 7.8, SD 0.6), ensuring comprehension across diverse literacy levels and supporting patient engagement in recovery planning.

### System Configuration

The system is engineered to align with the constraints of clinical workflows, enabling future testing and integration with existing imaging systems. It adheres to PACS communication protocols, receiving raw DICOM data and delivering DICOM-encapsulated reports that embed results into radiological workflows. Operating as a Docker container, to ensure stable behavior across platforms, the system is distributed through the Kubernetes platform to automatically scale and handle varying workloads, such as processing data from multiple patients. A locally hosted single LLM server, running in a separate Docker container, receives and processes only extracted features from the system, without accessing sensitive patient information, ensuring HIPAA compatibility. See Supplemental Materials for detailed methods.

## Results

Results are divided into separate sections describing the performance of the data processing pipeline and the performance of the cognitive outcome prediction models. In each case, the performance was evaluated on the test cohort of 160 patients with new-onset acute ischemic stroke with clinically acquired MRI scans, 153 of whom had data for any cognitive outcome measure. We report results from a single, fully automated processing run on the out-of-sample clinical test dataset.

**Data Selection, Processing and Registration.** All 160 ischemic stroke cases in the test dataset met minimum eligibility criteria, requiring at least one structural image, DWI and ADC) sequence. The pipeline successfully identified all DICOM sequences using an automated classification framework, transformed them into NIFTI format, and applied deep learning algorithms to compute brain masks and normalized images. Poor-quality data, such as images with insufficient brain coverage or artifacts, were automatically rejected based on objective quality metrics. This process resulted in all test cases being successfully registered to the MNI- 152 template, leveraging high-quality synthesized T1w images to enhance registration accuracy. Intermediate outputs, including image classifications, brain masks, and registrations, were visually reviewed to confirm quality across all cases.

**Lesion Segmentation.** Lesion segmentation performance was evaluated in three ways using a subset of 57 patients (N=57) from 160 cases with manually traced lesions, totaling 112 lesions. First, segmentation quality was evaluated with a Dice coefficient, achieving a mean of 0.69 (SD=0.21). For a more relevant, modern subset (data acquired post-2015), the mean Dice score improved to 0.74 (**Supplementary Tables 4–5**). Second, detection analysis was performed to test the model’s ability to identify individual lesions (N=112) beyond overall segmentation quality, with the model detecting 93% of lesions ≥ 1 cm³ and 98% of lesions ≥ 2.5 cm³. Third, to confirm that model-generated segmentations maintain predictive capability, we compared prognostic classification similarity between outcome prediction models using automated versus manual segmentations, yielding a 96% average correspondence across 681 cognitive outcome predictions (**Supplementary** Figure 6). This indicates that automated segmentations consistently capture behaviorally relevant anatomical features comparable to manual segmentations.

**Cognitive Outcome Prediction Results.** Classification performance for each neuropsychological outcome was evaluated using the AUC obtained by applying the trained model to the test dataset. AUC scores of 0.5 correspond to the performance that would be expected from a random classifier, and higher values reflect more accurate classification performance. Outcome prediction results from the independent test dataset are summarized in **Figure 4**, and results obtained from cross-validation analyses in the training dataset are shown in **Supplemental Figure 5.** AUCs varied substantially across the 28 outcomes evaluated in the training dataset. Many models achieved statistically significant classification performance as determined by permutation testing, with select models achieving excellent performance (i.e. AUCs >= 0.8), many models achieving intermediate performance (i.e. 0.6 < AUC < 0.8), and a few models performing worse than would be expected from a random classifier (i.e. AUCs < 0.5). Based on these results, we focused on five assessments that span clinically important functions with reasonably accurate predictions. This included expressive language, receptive language, visuospatial function, working memory, and executive function (described below, see **Figure 5**).

**Figure 4.**
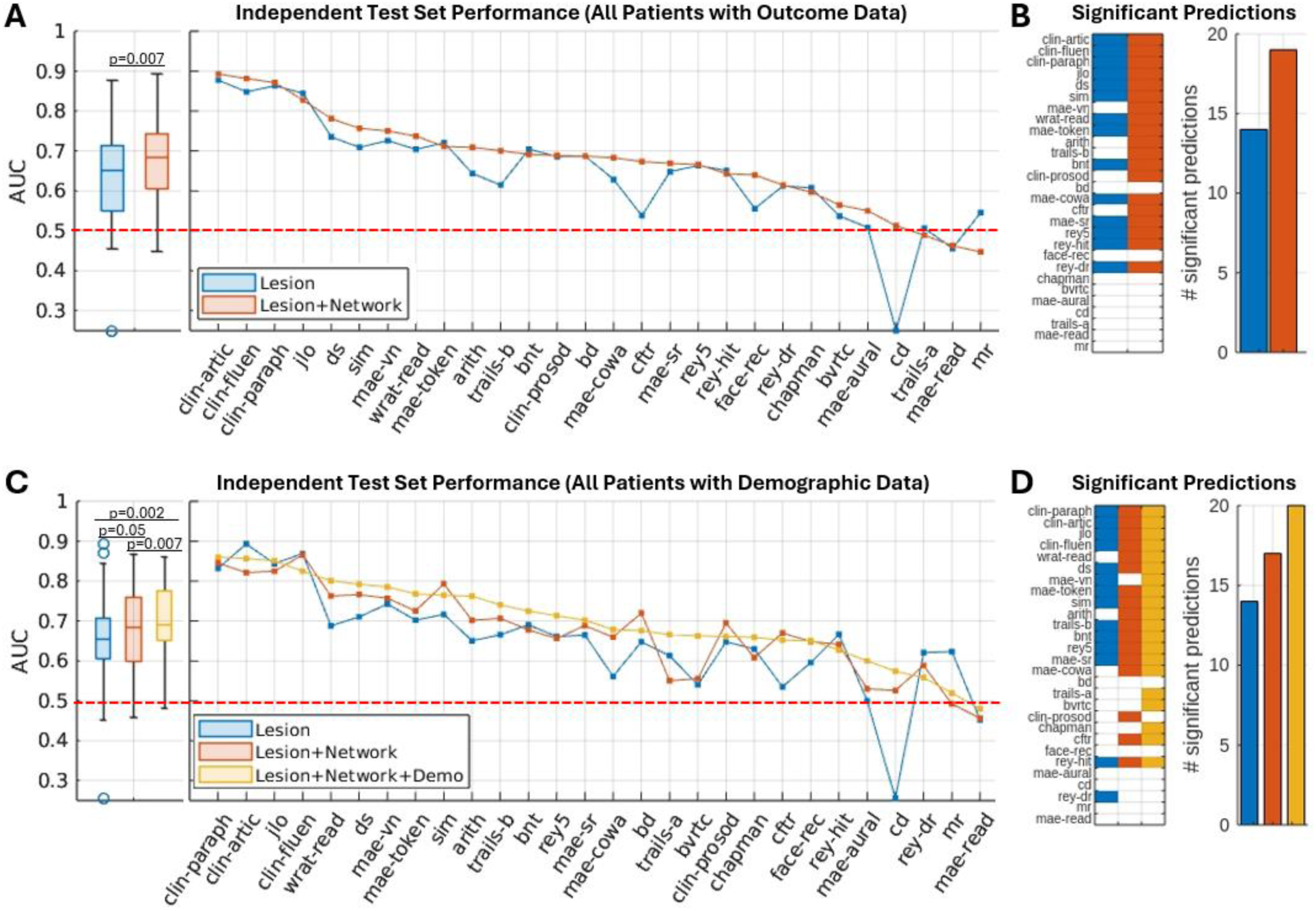
Cognitive outcome prediction results. **A.** Test dataset AUCs for the lesion location and stacked lesion + network models. The boxplots in the left panel show the distributions of AUCs across all outcome measures for each modeling strategy. The line plots in the right panel show AUCs for each outcome measure, sorted in descending order based on performance of the stacked lesion + network models. The dashed red lines correspond to chance performance levels that would be obtained by random guessing. **B.** The colored cells in the matrix correspond to statistically significant (p<0.05) test set outcome predictions as determined by permutation testing, while white cells indicate statistically non-significant (p>0.05) test set outcome predictions. The bar graph shows the total number of statistically significant outcome predictions for each modeling strategy. **C,D.** Same as (A,B), but for analyses evaluating whether adding demographic information (age, education) to the stacked lesion + network models can further increase performance. These analyses were restricted to the subset of patients with complete demographic data (36 training set patients excluded, 10 test set patients excluded).

**Figure 5.**
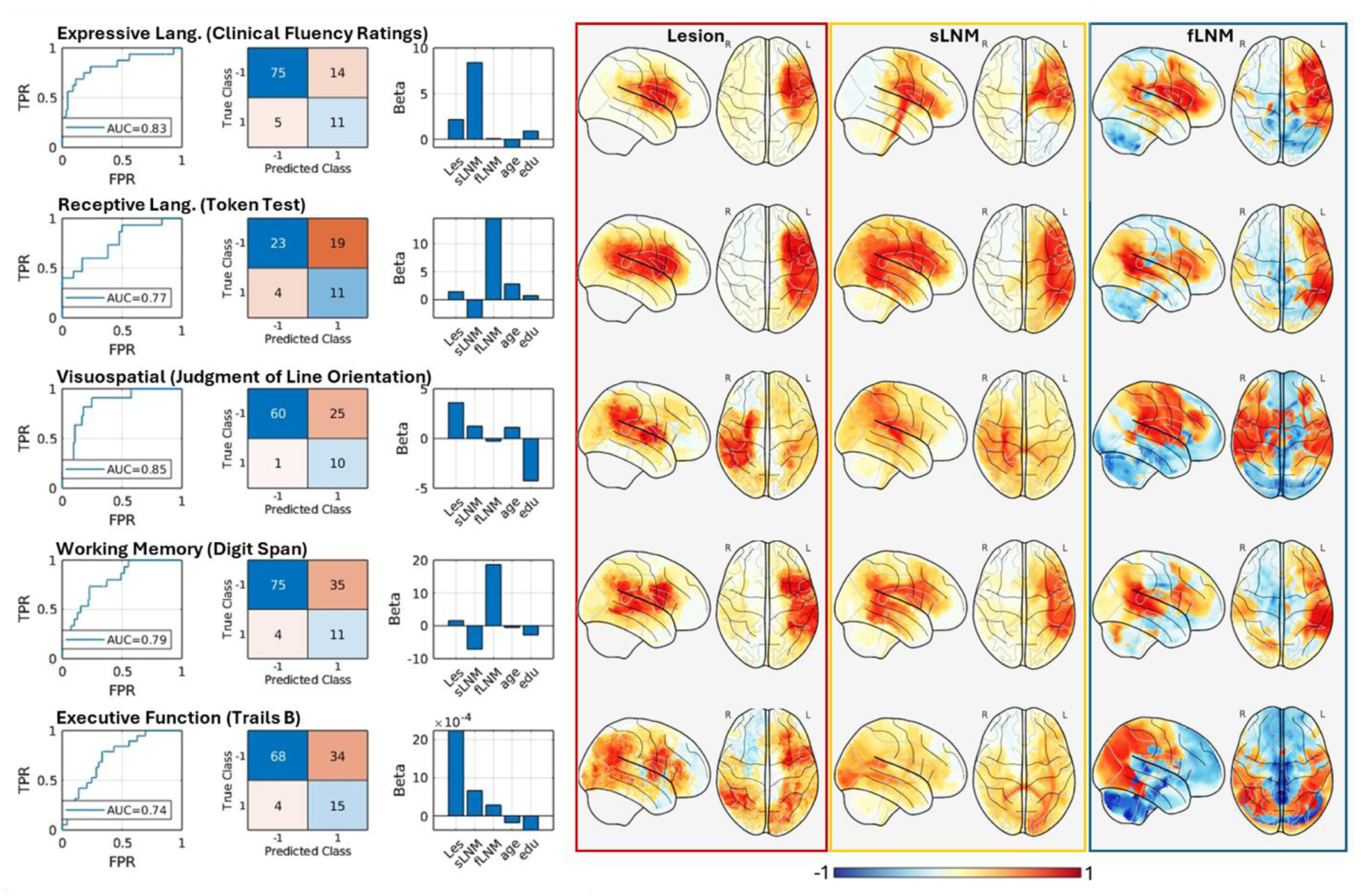
A detailed breakdown of 5 selected outcomes is shown in Figure 4C. Each row corresponds to a different outcome. The line plots show the ROC curve for each outcome, obtained by applying the trained model to the test dataset. The matrices show classification performance in the test dataset using the optimized classification score thresholds from the training dataset. The positive class corresponds to impaired patients, and the negative class corresponds to unimpaired patients. The bar plots show the model coefficients from the stacked logistic ridge regression models. Surface plots show the full (unthresholded) voxel-wise PLS classification model coefficients from the individual lesion location, fLNM, and sLNM models. PLS classification model weights were normalized to the range [-1, 1] by dividing all weights in each map by the maximum absolute weight value for visualization purposes.

In line with prior work, models that incorporated both lesion location and lesion network information often performed better than models that relied on lesion location alone **(Figure 4, A- B).** Across outcomes, models incorporating lesion and network information achieved significantly higher AUC values than models relying on lesion location alone (signed rank test, p=0.007). The lesion + network models also achieved more statistically significant (i.e., permutation p<0.05) classification results in the test dataset than the lesion-only models (**Figure 4C**), indicating that the addition of network information enabled more robust classification performance. Adding demographic information (age, education) further improved performance **(Figure 4, C-D).** Models incorporating lesion, network, and demographic information achieved significantly higher AUC values across outcomes than both the lesion + network models (signed rank test, p=0.007) and the lesion-only models (signed rank test, p=0.002), and achieved more statistically significant (permutation p<0.05) outcome predictions (**Figure 4D**). These results demonstrate that incorporating lesion-derived network information and demographic data can enhance cognitive outcome prediction beyond what is afforded by lesion location alone.

Full names of each outcome are provided in the **Supplementary Material**. Note – for panels (B,D), the outcomes on the y-axes are ordered the same way as those on the x-axes in panels (A,C).

For descriptive purposes, **Figure 5** shows detailed characterizations for 5 models targeting distinct cognitive outcomes spanning expressive language (clinical fluency ratings), receptive language (Token Test), visuospatial (Judgment of Line Orientation), auditory working memory (Digit Span), and executive function (Trails B) domains. In general, each of these models performed well in correctly classifying impaired patients in the test dataset (i.e., sensitivity) at the trained classification score thresholds, but they exhibited more variable performance for correctly classifying unimpaired patients (i.e., specificity). Some models, such as the model for Judgment of Line Orientation, achieved good performance for both groups. This model correctly classified 91% of impaired patients and 71% of unimpaired patients, demonstrating good sensitivity and specificity. Other models, however, demonstrated good sensitivity but poor specificity. For example, the model for the Token Test correctly classified 73% of impaired patients but only correctly classified 55% of unimpaired patients. In the context of the moderate-to-high AUCs for these models, which indicate that the predicted classification scores can achieve robust group discrimination across classification score thresholds, these observations suggest that the optimized classification score thresholds learned from the training dataset may not always be optimal in the test dataset even if the models themselves have good predictive power. This could reflect differences between the training and test datasets in terms of lesion etiology, lesion segmentation methods (automated vs. manual), and/or lesion chronicity.

Improving the generalizability of trained classification score thresholds is an important goal for future iterations of the cognitive outcome prediction models.

Model weight patterns varied substantially across the 5 models shown in **Figure 5**, highlighting that the predictive value of different features varies depending on the predicted outcome. The coefficient maps from the individual models highlight variations in the predictive anatomy within domains (e.g., expressive vs. receptive language outcomes) and between domains (e.g. language vs. visuospatial). Perhaps surprisingly, they also highlight similarities between domains and suggest a partially overlapping predictive anatomy between Token Test (i.e., receptive language) and Digit Span (i.e., auditory working memory), suggesting that deficits in receptive language and deficits in auditory working memory may result from damage to the same or partially overlapping brain systems.

**Report Generation.** The automated generation of reports performed as expected, with several examples included in the supplemental material. A clinical neurologist reviewed randomly selected reports and confirmed they provided supportive information with potential clinical utility. The neurologist noted that the reports contained no errors that could lead to patient harm. The neurologist expressed enthusiasm for the reports’ potential in a clinical setting, supporting their value as a proof-of-concept. Further validation is needed before clinical implementation.

**Runtime Analysis.** Runtime performance was assessed on a workstation equipped with an Intel Xeon w7-3545 CPU (24 cores, 48 threads, 4.8GHz max frequency) and an NVIDIA RTX 6000 Ada GPU (48GB memory). The system achieved an average end-to-end processing time of 121.3 seconds (approximately 2 minutes), with 95% of cases completing in under 3 minutes (range=80.4 to 238.1 seconds, SD=22.3), as shown in **Table 1**. Key pipeline stages, including data selection, registration, lesion segmentation, outcome prediction, and report generation, were optimized for efficiency, with report generation benefiting significantly from GPU acceleration of large language model inference. This rapid processing speed supports the system’s feasibility for acute stroke care settings, where timely analysis is critical. Detailed performance benchmarks, including CPU-only runtimes and individual component breakdowns, are provided in **Supplementary Table 8.**

**Table. 1.** Runtime summary of processing system components in seconds.

| System Component | Mean | Min | Max | Std |
| --- | --- | --- | --- | --- |
| Data Selection and Preprocessing | 29.6 | 18.8 | 48.0 | 5.0 |
| Registration | 76.7 | 49.1 | 168.8 | 15.4 |
| Lesion Segmentation | 4.1 | 3.9 | 4.6 | 0.2 |
| Outcome Prediction | 0.9 | 0.8 | 0.9 | 0.0 |
| Report Generation | 10.0 | 7.8 | 15.8 | 1.65 |
| Total | 121.3 | 80.4 | 238.1 | 22.3 |

## Discussion

Here, we demonstrate for the first time a clinical neuroimaging platform for fully automated data processing and outcome prediction in patients presenting with acute ischemic stroke.

Importantly, our platform can generate personalized outcome predictions from raw clinical imaging data in less than 3 minutes and communicate them to patients and clinicians in automatically generated reports. We demonstrated robust performance of the automated processing pipeline, with all 160 MRIs from the test dataset being processed without any failures. The deep learning algorithm for stroke segmentation performed adequately, with excellent performance for lesions >1 cm^3^. We also demonstrated promising out-of-sample prediction performance across a range of neuropsychological outcome measures. We showed that incorporating lesion-derived network information and demographic variables can enhance prediction performance (**Figure 4**) and highlighted performance for a set of outcome measures spanning distinct cognitive domains (**Figure 5**). This work represents an important advance towards harnessing the translational potential of lesion research for informing stroke prognostication.

Historically, lesion-symptom mapping and related fields have primarily been concerned with identifying anatomical structures that show statistically significant group-level associations with lesion location ^54,55^. However, the pursuit of personalized, precision medicine approaches to stroke prognostication requires the development of lesion-symptom models that can reliably predict outcomes in individual patients ^56^. Furthermore, for such models to be clinically useful, they must be able to generalize beyond high-quality research datasets to real-world clinical data, and they must be implemented in a fully automated platform that generates outcome predictions from raw clinical imaging quickly and easily. Our approach, which embeds pre-trained brain-behavior models within a fully automated end-to-end clinical data processing pipeline, solves these issues and represents a major step towards developing imaging-based clinical tools for ischemic stroke outcome prognostication. Notably, excellent classification performance was consistently achieved for measures corresponding to clinician ratings of impairment in fluency, articulation, and paraphasias (**Figure 4**, “clin-fluen”, “clin-artic”, “clin- paraph”), indicating that brain-behavior models can robustly predict chronic “real-world” clinical outcomes in a way that aligns with clinician evaluations.

While our brain-behavior models were trained on manual lesion segmentations (and derived network features) from research datasets, they nonetheless were able to achieve meaningful and statistically significant outcome prediction performance when applied to automatically generated lesion segmentations (and derived network features) across a variety of cognitive outcome measures, including measures of distinct cognitive domains, in a fully independent clinical test dataset (**Figures 4-5**). This is a strong proof-of-concept for our approach, demonstrating that lesion information derived from clinical scans can be successfully leveraged to predict chronic cognitive outcomes in individual patients. While from a machine learning perspective, it would be ideal to train brain-behavior models on acute lesion segmentations produced by the automated pipeline (i.e. to maximize the representativeness of the training dataset with respect to the data that models will be applied to in practice), most existing lesion-symptom mapping datasets consist of manual lesion segmentations from chronic research-quality structural scans. Importantly, our results suggest that models trained on manual lesion segmentations from research datasets can still produce meaningful predictions when applied to automatically generated lesion segmentations from acute clinical scans, suggesting that large existing research datasets can be leveraged to develop brain-behavior models to predict outcomes in acute ischemic stroke.

Notably, our results demonstrate that even relatively simple measures of lesion-network properties and patient demographics can yield significant and sometimes drastic improvements in prediction performance beyond what is possible with lesion location (**Figure 4**). We anticipate continued improvements in prediction performance as future predictive models expand on the current range of both imaging and non-imaging predictors. Lesion network mapping approaches will likely continue to improve, along with the incorporation of other imaging variables with predictive utility for long-term functional outcomes, like quantifying white matter hyperintensity ^57^, large vessel occlusion ^58^, cortical atrophy ^59^, and brain age ^60^. Tissue segmentation and normative modeling could also be leveraged to improve predictive models ^61^. With new training datasets, it will be possible to expand the range of clinical and demographic variables included in the statistical models, such as medical co-morbidities, more accurate estimation of pre-stroke baseline, social support estimates, and acute impairment measures, such as the NIH stroke scale. It may be possible for these non-imaging variables to be extracted from electronic medical records or entered manually. The ongoing collection of longitudinal outcomes from large and well-characterized stroke cohorts (e.g. DISCOVERY, VERIFY) will facilitate the development of these expanded models ^62,63^.

Lastly, it is essential to note that we are not advocating for the clinical use of this tool for outcome prediction in its current form. Instead, we view the current overall performance and proof-of-concept analysis as a promising starting point for further development. There are many limitations of the current approach that can be improved upon. For example, our deep learning algorithm for lesion segmentation has improved consistently as training data has been added, and the current 700 MRIs are likely not approaching peak lesion segmentation accuracy.

Further, our training dataset may be suboptimal as a mixed lesion etiology cohort rather than a stroke-specific cohort, especially given the very low rates of impairment for many outcomes; larger training datasets with better representation of impaired patients would be expected to improve prediction performance. Relatedly, individual cognitive/behavioral tests may not be ideal targets for eventual clinical applications. The development and refinement of models that can predict clinically relevant outcomes (e.g., expressive language, visuospatial function, motor function) instead of performance on specific neuropsychological tests is an important future direction for the pipeline that will likely require the harmonization of within-domain outcome data across different assessments and likely across multiple independent datasets.

In closing, this paper presents the first fully automated clinical neuroimaging platform for lesion-informed prognostication of ischemic stroke. With continued development, we are optimistic that this approach can extract clinically valuable information from routinely acquired clinical neuroimaging and put it in the hands of clinicians and patients, forming a much-needed bridge connecting advances in lesion research to improvements in precision medicine.

## Data Availability

The software used for lesion modeling can be downloaded at the provided URL. Deidentified data from this study may be made available upon reasonable request to the corresponding author, subject to a data use agreement ensuring compliance with human subjects ethical standards. Trained machine learning models for outcome prediction may be shared, subject to a proper software use agreement protecting intellectual property rights.

## Acknowledgments

The authors wish to thank the patients with stroke who participated in this research. Dan Tranel and Ken Manzel helped with data collection. Mark Bowren helped with the selection, organization, and norming of cognitive outcome data and defining impairment thresholds.

## Author Contributions

M.B. designed and implemented the data processing approaches, conducted the data analysis and creation of figures related to processing pipeline development, and drafted the manuscript.

J.C.G. designed and implemented the cognitive outcome modeling approaches, performed the data analysis related to cognitive outcome prediction, created figures related to outcome prediction, and drafted the manuscript. J.B. contributed to data preparation and lesion tracing and supported analyses. C.S. contributed to behavioral and demographic data organization.

C.J.R. contributed to the pipeline’s report generation, testing, and engineering. H.J. supervised the engineering aspects of the pipeline development. A.D.B. contributed to the conceptualization, design, and supervision of the study. All authors revised and approved the final version of the manuscript.

## Supplementary Material

See Supplemental File.

