## Supplemental Materials for "A Clinical Neuroimaging Platform for Rapid, Automated Lesion Detection and Personalized Post-Stroke Outcome Prediction"

Supplementary Materials

**Automated Image Processing Pipeline**

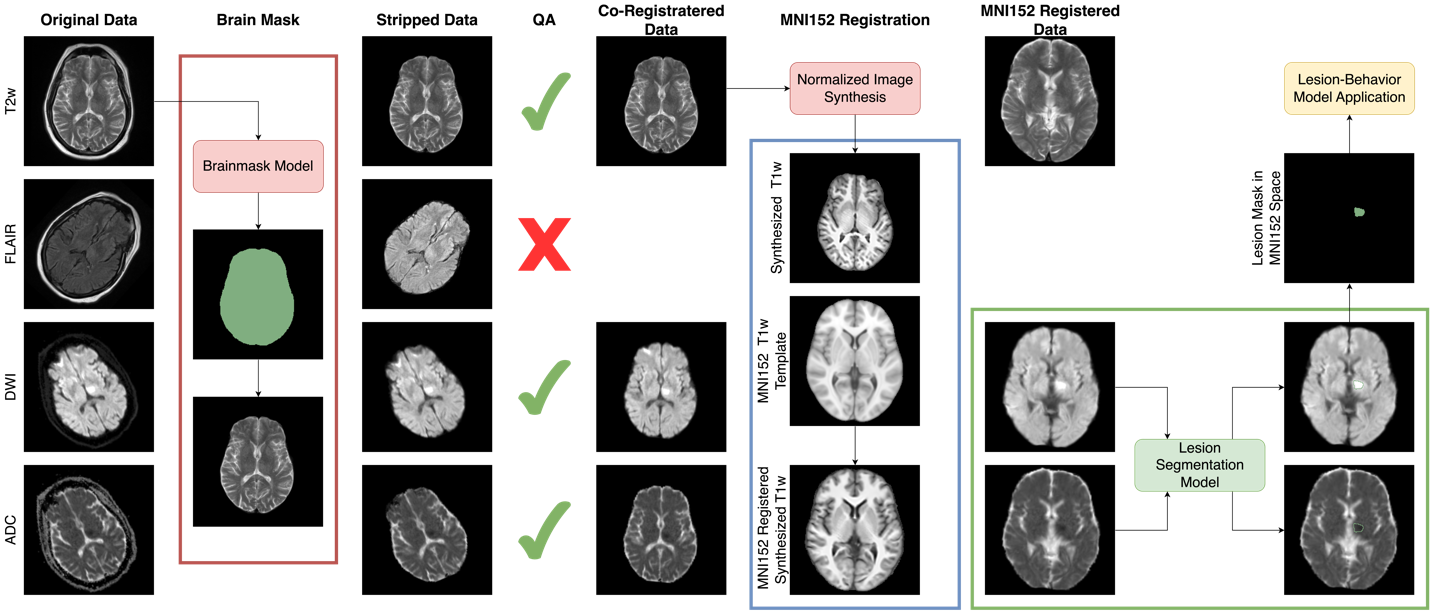

**Supplemental Figure 1.** Visualization of brain MRI processing steps from raw clinical image to segmented lesion masks in MNI152 template space for lesion-behavior model application.

***Development Datasets***

**Iowa Dataset.** For the development and validation of the automated processing pipeline, we utilized a retrospective dataset of 1,047 ischemic stroke patients from the University of Iowa Hospital and Clinics (UIHC) archives (IRB-approved and anonymized). This dataset comprised standard-of-care MRI scans acquired within one week of stroke onset, spanning from December 2009 to October 2021. The imaging data exhibited substantial technical heterogeneity, having been collected across 20 unique scanner models from 5 major manufacturers (Siemens, Philips, GE, Toshiba, and Hitachi). This diversity of acquisition sources intentionally introduced technical variability that strengthened the generalizability of our pipeline to heterogeneous clinical environments.

Different subsets of this dataset were strategically employed for various pipeline components: fine-tuning the DICOM classifier tool, developing and validating brain mask models, training lesion segmentation algorithms, and optimizing registration approaches. The extensive temporal range and diversity of acquisition parameters enhanced the robustness of our models to clinical variability.

**ISLES 2022 Dataset.** The ISLES 2022 dataset was obtained from the Ischemic Stroke Lesion Segmentation Challenge (ISLES22) ^1^ and contains 250 subjects with scans of varying quality. This dataset was designed specifically for the evaluation of automated stroke lesion segmentation methods and represents a multi-center collection of clinical stroke MRI data. The challenge dataset includes multi-modal MRI sequences (DWI, ADC, FLAIR) acquired in acute stroke settings across multiple international medical centers, capturing a wide range of lesion sizes, locations, and imaging protocols. Unlike curated research datasets, ISLES 2022 intentionally incorporates the real-world variability found in clinical environments, including motion artifacts, varying field strengths, and diverse acquisition parameters. This heterogeneity makes it particularly valuable for validating the robustness and generalizability of automated stroke processing pipelines in realistic clinical scenarios.

*Brain Mask Model*

The brain mask model training dataset was derived from the Iowa dataset described above. As depicted in Supplementary Table 1, we utilized 4,823 images from 1,047 patients across five MRI modalities: T1-weighted (T1w), T2-weighted (T2w), Fluid-Attenuated Inversion Recovery (FLAIR), and Diffusion Weighted Imaging derivatives—Apparent Diffusivity Coefficient (ADC) and Diffusion Trace (DWI). We employed a three-fold approach to model development: initial model training with 3,831 images from 837 subjects, hyperparameter optimization with an independent validation set of 496 images from 104 subjects, and final performance assessment on a separate test set of 496 images from 106 subjects. This corresponded to an approximate 80:10:10 percent split at the subject level, ensuring that images from the same subject never appeared across different datasets to prevent data leakage.

The Ground Truth masks used for training the brainmask model were initially generated using the SynthStrip algorithm. Each mask was then manually verified and updated where needed to ensure accuracy. While SynthStrip generally performs well, we identified failure cases, particularly with diffusion images. Our custom model is specifically trained on stroke clinical data with extensive data augmentation, resulting in robust performance fine-tuned for stroke clinical imaging and the intended purpose of this system. This approach gives us full control over the pre- and post-processing steps, model files, and enables efficient integration within the system.

**Supplementary Table 1. Brain Mask Datasets**

| Modality | Training | Validation | Testing | Total |
| --- | --- | --- | --- | --- |
| T1w | 754 | 101 | 98 | 958 |
| T2w | 577 | 79 | 85 | 741 |
| ADC | 839 | 106 | 106 | 1051 |
| Flair | 817 | 104 | 101 | 1022 |
| DWI | 839 | 106 | 106 | 1051 |
|  | = 3831 | = 496 | = 496 | = 4283 |

The automated brain extraction used 3D ResUNet model (Kerfoot et al., 2019) demonstrated remarkable consistency across multiple MRI modalities. We used MONAI implementation of the model and trained using PyTorch and PyTorch-Lightning libraries ^2^. We evaluated the model on five different sequences commonly used in stroke imaging: T1-weighted, T2-weighted, FLAIR, ADC and DWI images. On the test dataset, the model achieved a mean DICE score of 0.98 across all modalities, with notably low variability (standard deviation ≤ 0.01 for all sequences). Boundary accuracy, measured by balanced Average Hausdorff Distance i.e. bAVD ^3^, was also highly consistent, ranging from 0.16 to 0.24 voxels across modalities. This stability across different imaging contrasts is particularly important for robust preprocessing in clinical settings where multiple MRI sequences are routinely acquired.

**Supplementary Table 2. Brain Mask Results**

| Modality | DICE | | bAVD | |
| --- | --- | --- | --- | --- |
|  | Mean | Std | Mean | Std |
| T1w | 0.98 | 0.00 | 0.16 | 0.00 |
| T2w | 0.98 | 0.00 | 0.16 | 0.00 |
| ADC | 0.98 | 0.01 | 0.24 | 0.01 |
| Flair | 0.98 | 0.01 | 0.17 | 0.01 |
| DWI | 0.98 | 0.00 | 0.16 | 0.01 |

#### *Registration*

**Methods.** Registration in clinical stroke imaging serves multiple critical purposes. Co-registration aligns images from the same session to correct for patient movement and inherent acquisition differences between modalities, while template registration enables standardized anatomical analysis through brain atlases and application of pre-existing statistical models for outcome prediction. The presence of stroke lesions presents unique challenges for registration, particularly when aligning pathological brains to healthy templates. While traditional approaches often employ explicit lesion masking or enantiomorphic correction, these methods require accurate lesion segmentation prior to registration and have limitations with lesions crossing the midline or in cases of asymmetric brain structure.

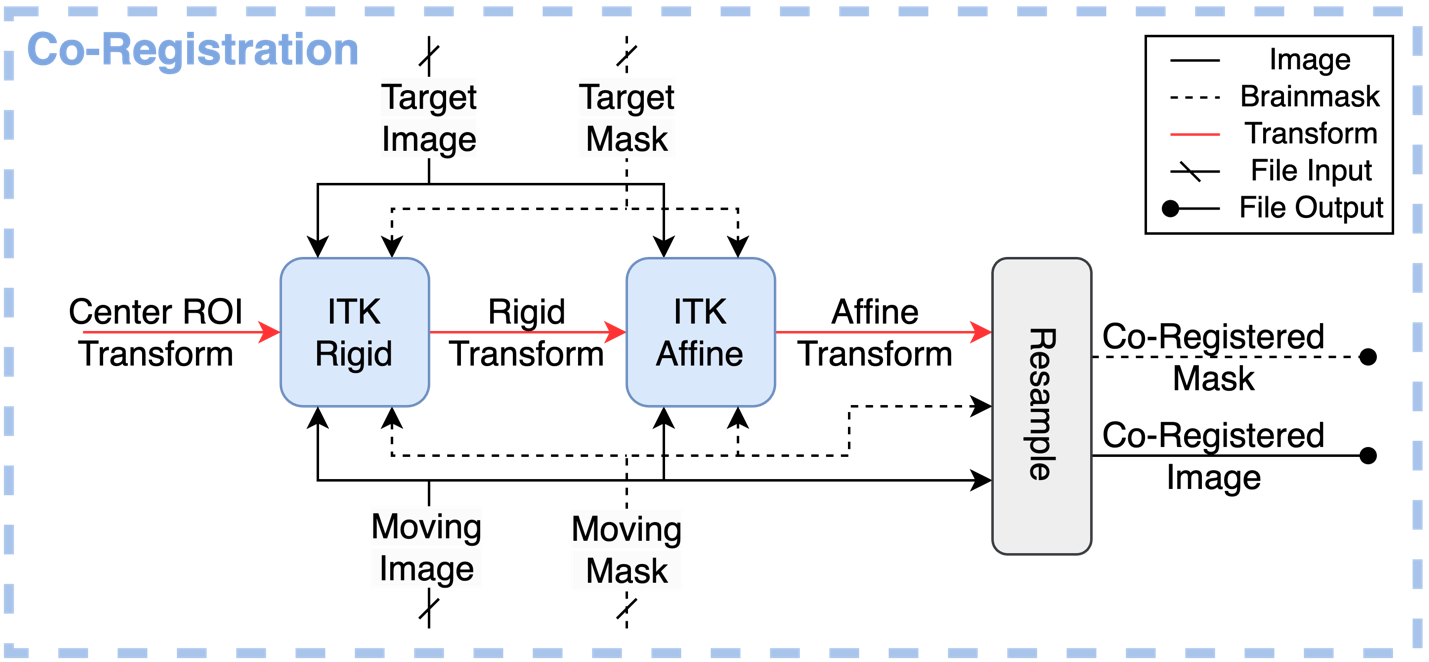

**Supplementary Figure 2.** Co-registration pipeline utilizing sequential rigid and affine registration steps. The process employs brain masks to guide registration metrics, with the target image serving as the fixed reference while remaining modalities are treated as moving images.

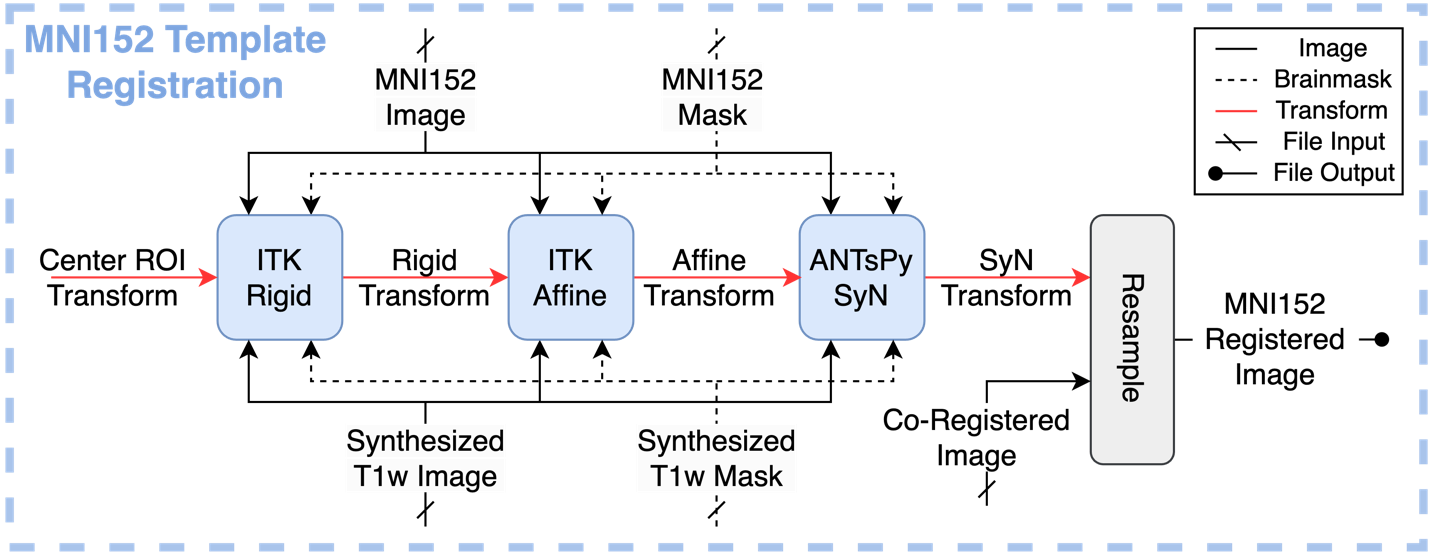

**Supplementary Figure 3.** Simplified View of the Template Registration module showing the three-stage registration process that transforms patient-specific MRI data into standardized MNI152 space.

Our registration framework addresses these challenges through a comprehensive workflow that first co-registers all modalities to the highest quality anatomical image selected during preprocessing. This process employs sequential rigid and affine transformations implemented through ITK, with brain masks defining the registration sampling region to enhance accuracy, following methodology in BRAINSFit ^4^. For template space registration, we utilize a three-stage approach (rigid, affine, and SyN deformable registration) to align the synthesized T1w image with the MNI-152 template. Rather than explicitly handling lesions, our approach leverages SynthSR-generated T1w images that normalize tissue contrast and effectively manage lesioned areas through implicit inpainting while maintaining anatomical consistency. This synthetic approach works effectively across both acute and chronic stroke phases without requiring separate lesion segmentation, dramatically simplifying the clinical workflow. The final stage applies the resulting transformation to all co-registered patient images, ensuring consistent spatial normalization across all modalities while minimizing interpolation artifacts.

**Results.** We validated our registration approach on a comprehensive dataset of 2,987 clinical MRI scans derived from the Iowa dataset and comprising 1,122 FLAIR images (37.6%), 1,041 T1-weighted images (34.9%), and 824 T2-weighted images (27.6%) from diverse acquisition protocols, scanner manufacturers, and field strengths. To assess the effectiveness of our synthetic image-based approach, we conducted a comparative analysis between conventional registration using original clinical images and our synthetic pipeline using SynthSR-generated T1w images. Both pipelines employed identical registration parameters, with evaluation metrics calculated on the original clinical images after transformation to ensure fair comparison.

The synthetic image-based approach demonstrated clear advantages in both accuracy and robustness. The synthetic approach achieved consistently higher DICE scores between registered brain masks and the MNI template mask (0.960 vs. 0.942) across all imaging modalities. Most notably, while the conventional pipeline resulted in 44 failures out of 2,987 cases (98.5% success rate), the synthetic pipeline reduced this to only 8 failures (99.7% success rate). This exceptional reliability has significant implications for clinical deployment, minimizing the need for time-consuming manual corrections while enabling successful processing of cases that would typically fail in conventional pipelines. The synthetic approach's consistent performance across varying input data quality is particularly valuable in clinical settings where imaging protocols may vary considerably, confirming its suitability for routine clinical deployment.

### *Lesion Segmentation*

**Model and Image Processing.** Our automated lesion segmentation approach utilizes both Apparent Diffusion Coefficient (ADC) and DWI images as complementary input modalities. The preprocessing pipeline, detailed in our previous work ^5^, includes intensity normalization using percentile-based scaling (-1 to 1) computed from foreground intensities, while deliberately avoiding hard intensity clipping to preserve the distinction between true lesions and hyperintensities. All images are standardized to 1mm isotropic spacing with consistent dimensions (192×192×160), using center-of-gravity-based resampling to ensure standardized input for GPU processing.

The model architecture employs a 3D Residual U-Net design ^6^, combining the semantic segmentation capabilities of 3D U-Net ^7^ with the training advantages of residual connections. This architecture incorporates an encoder-decoder pathway with skip connections, enabling detailed localization while preserving contextual information. The network utilizes strided convolutions for downsampling and transpose convolutions for upsampling and includes residual units in both encoder and decoder paths to address the vanishing gradient problem.

Post-processing includes converting probability maps to binary segmentation masks followed by in-plane morphological operations (2×2 voxel dilation followed by 1×1 erosion) to refine segmentation boundaries while maintaining anatomical coherence. These operations are applied only in-plane to preserve the original slice-wise characteristics of the segmentation. All pre and post processing methods were implemented using ITK and MONAI libraries.

**Model Training.** The model was trained on a combined dataset comprising approximately 450 subjects from the Iowa dataset with manually delineated lesion masks and 250 subjects from the ISLES2022 dataset. Training was performed on an NVIDIA RTX8000 GPU with 48GB of memory. While all ISLES2022 subjects were included in the training dataset, the Iowa dataset was split into training, validation, and test sets. The validation set was used during the training process to monitor performance and select the optimal model checkpoint, following standard deep learning practices. The final model achieved an average DICE coefficient of 0.74 on the held-out test dataset containing 100 subjects. This trained model was subsequently implemented in the system and evaluated on a separate clinical test dataset, as detailed in later sections.

**Input Modality Analysis.** To determine the optimal input modality combination, we conducted a comparative analysis of different MRI sequence combinations for lesion segmentation. Four identical 3D Residual U-Net models were trained using the same subset of the Iowa Stroke Retrospective Dataset (342 subjects containing all five modalities of interest: DWI, ADC, T1w, T2w, and FLAIR), with consistent data splits across all configurations (80% training, 10% validation, 10% testing). These models varied only in their input modalities: (1) DWI+ADC only, (2) DWI+ADC+T1w, (3) DWI+ADC+T2w, and (4) DWI+ADC+FLAIR. As shown in Supplementary Table 5, adding anatomical sequences provided no statistically significant improvement over diffusion-weighted images alone. The baseline model (DWI+ADC) achieved a mean DICE score of 0.792, comparable to models incorporating additional anatomical sequences (DICE 0.788-0.799). This finding aligns with clinical understanding that acute stroke lesions are often not visible on structural images, being primarily detectable on diffusion-weighted sequences. The absence of meaningful performance gains, coupled with considerations of data availability, computational efficiency, and model stability, strongly supported our decision to use only diffusion-weighted sequences in the final implementation. This streamlined two-modality approach offers practical advantages including reduced preprocessing time, smaller model footprint, and improved system efficiency without sacrificing segmentation accuracy.

**Supplementary Table 3. Lesion Segmentation Input Modalities Analysis**

| **Input Modalities** | **DICE Mean** | **DICE Std** | **bAVD Mean** | **BAVD Std** |
| --- | --- | --- | --- | --- |
| DWI + ADC (Baseline) | 0.79 | 0.2 | 0.70 | 1.30 |
| DWI + ADC + T1w | 0.80 | 0.2 | 0.92 | 3.02 |
| DWI + ADC + T2w | 0.79 | 0.20 | 0.58 | 1.08 |
| DWI + ADC + FLAIR | 0.79 | 0.21 | 3.03 | 9.60 |

**Test Dataset Segmentation Results.** The automated lesion segmentation model was evaluated on an independent clinical test dataset comprising 57 subjects with expert-validated lesion labels. As shown in Table 4. the model achieved a mean DICE score of 0.69 across all cases, with a standard deviation of 0.21. After excluding two failed cases, performance improved to a mean DICE of 0.71 with reduced variability (standard deviation of 0.16). The balanced Average Hausdorff Distance (bAVD) of 0.95 voxels indicated strong boundary agreement between predicted and ground truth segmentations.

**Supplementary Table 4. Lesion Segmentation Results**

| **Metric** | **DICE** | **bAVD** |
| --- | --- | --- |
| Mean | 0.69 | 0.95 |
| Std | 0.21 | 1.48 |

**Temporal analysis.** Temporal analysis of model performance revealed a clear trend correlated with image acquisition periods, with results shown in Table 5. The dataset spanned over two decades (2001-2024), allowing for evaluation across different eras of MRI technology. The model achieved a DICE score of 0.74 on recent data (post-2015), compared to 0.68 for cases from 2011-2015 and 0.66 for pre-2011 cases. This improvement is particularly noteworthy given that recent cases had smaller average lesion volumes (47.6 cm³ post-2015 vs 54.1 cm³ pre-2011), as DICE scores typically favor larger lesions (Raina et al., 2023). The consistent improvement across time periods, achieved despite this inherent DICE bias, suggests that the model effectively leverages the enhanced image quality of modern MRI scanners.

**Supplementary Table 5. Lesion Segmentation Results by Time Period**

| **Period** | **n** | **Avg. Lesion Vol. (cm³)** | **DICE** |
| --- | --- | --- | --- |
| Before 2011 | 9 | 54.1 | 0.66 |
| 2011-2015 | 39 | 40.5 | 0.68 |
| After 2015 | 9 | 47.6 | 0.74 |

**Lesion Detection Analysis.** We conducted a lesion-wise detection analysis on our test dataset comprising 57 subjects with 112 individual lesions. For clarity, each connected component (individual island) in the binary lesion mask was counted as a separate lesion, which explains the multiple lesions per patient. Successful detection was defined as any overlap between the model's prediction and the ground truth lesion mask. The model demonstrated an overall detection accuracy of 0.81 across all lesions. Excluding the small lesions under 1 cm³, accuracy improved markedly to 0.93 for lesions ≥ 1 cm³ and 0.98 for lesions ≥ 2.5 cm³. When excluding two previously identified failed cases (analyzing 106 lesions across 55 subjects), detection rates further improved to 0.88, 0.98, and 1.00 respectively for all lesions, lesions ≥ 1 cm³, and lesions ≥ 2.5 cm³.

***Report Generation***

**Advanced Radiological Visualization Methods**

Our radiological visualization component incorporates multimodal 2D clinical imaging and enhanced 3D rendering techniques. For 2D visualization, the system automatically generates a standardized three-panel clinical view comprising Diffusion-Weighted Imaging (DWI), Apparent Diffusion Coefficient (ADC), and anatomical reference images. Each modality provides complementary insights: DWI highlights acute ischemic changes through hyperintense signal, ADC confirms true diffusion restriction through hypointense signal, and the anatomical image provides structural context. To enhance lesion visibility, we select slice with the highest lesion area and highlight the lesion boundary in red. For 3D representation, we employ the Visualization Toolkit (VTK) to create spatial visualizations in standardized MNI space, ensuring consistency across reports. Our rendering pipeline utilizes ray-casting volume rendering with optimized transfer functions to generate semi-transparent brain templates with lesion overlays. The system automatically generates standard views in axial, coronal, and sagittal. This comprehensive visualization approach provides both clinicians and patients with complementary perspectives: 2D slices offering precise anatomical localization and signal characteristics, while 3D visualizations reveal the full spatial extent and distribution of damage.

**Neuroanatomical Analysis Framework**

Our neuroanatomical analysis framework integrates two complementary brain atlases to provide comprehensive characterization of stroke lesions: an arterial territory atlas defining 30 vascular regions and an anatomical atlas parcellating the brain into 154 distinct regions. The anatomical structures atlas builds upon the Harvard-Oxford Cortical Atlas ^8,9^, enhanced through integration of subcortical structures from the Harvard-Oxford Subcortical Atlas ^10,11^. We incorporated the Morel Thalamic Atlas ^12^ for more detailed thalamic regions, with careful boundary adjustments to maintain anatomical consistency. Brainstem regions were derived using FreeSurfer v6 processing of the MNI152 template ^13^, while cerebellar parcellation utilized the SUIT atlas ^14,15^. White matter regions, including corpus callosum and cerebral/cerebellar white matter territories were defined using a population-level tractography atlas ^16^. The vascular territory mapping was incorporated from Liu's arterial supply atlas ^17^, modified to create a hierarchical organization where broad arterial territories were derived from detailed vessel-specific regions, with ventricle regions excluded from label maps for improved accuracy. Using inverse transformation matrices computed during the registration pipeline, these atlases are warped into each patient's native image space using affine and deformable transformations, ensuring accurate regional analysis while preserving individual neuroanatomical variability.

After atlas transformation to patient space, our system computes detailed volumetric overlap statistics between the segmented lesion and each atlas region utilizing a voxel-wise approach. This quantitative analysis captures three critical measurements for each affected structure: absolute lesion volume in cubic centimeters (calculated by multiplying the voxel count by the voxel dimensions), percentage of total lesion volume within each region (volumetric proportion analysis), and percentage of each anatomical region affected by the lesion (region-specific burden assessment). The system employs binary masks for precise calculation of spatial overlaps, with partial volume effects addressed through proportional allocation methods. Results are presented in a structured, hierarchical format organized across three levels: hemisphere, cortical anatomical regions (lobes or arterial territories), and subcortical structures. This multi-tiered organization enables rapid assessment of both macroscopic damage distribution and detailed local impact assessment. The system's white matter versus gray matter analysis provides crucial context for understanding disconnection syndromes and potential functional implications, while the arterial territory analysis offers insights into the vascular etiology of the stroke, potentially informing secondary prevention strategies.

**Readability Analysis of Patient Communication**

We conducted systematic readability analysis of our patient-directed content using the Simple Measure of Gobbledygook (SMOG) formula ^18^, specifically chosen for its established use in evaluating health education materials. The SMOG index estimates the years of education needed to understand text by analyzing polysyllabic words within a sample of 30 sentences, providing a validated grade-level assessment. We randomly selected 100 subjects from our test set and analyzed their automatically generated patient communication sections, which are organized into four strategic sections: "What Happened," "What to Expect," "Your Risks," and "What to Do." These sections translate complex medical information into accessible language using relatable analogies and clear explanations, deliberately avoiding medical jargon. The SMOG analysis demonstrated consistent achievement of appropriate reading levels with a mean grade level of 6.6 (SD=0.6), maximum of 7.8, minimum of 4.9, and 95% confidence interval of 6.5-6.7. These results confirm that our patient-focused content consistently performs below the average US adult reading level (approximately 7th-8th grade), making the information accessible regardless of educational background and addressing the documented health literacy challenges faced by approximately 36% of US adults with basic or below basic health literacy skills ^19^.

**Technical Infrastructure and Clinical Integration**

Our report generation system employs a modular architecture built on ReportLab (version 3.6.0), an open-source Python library for programmatic PDF creation. The architecture follows a hierarchical structure where reports contain pages, and pages contain sections, enabling efficient management of report elements while maintaining formatting consistency. For narrative content generation, we employ Large Language Models (LLMs) using Ollama, an open-source LLM server framework deployed within a Docker container architecture. This deployment enables continuous model availability through a persistent server instance, with a containerized server maintaining the LLM in memory. The implementation employs Meta's LLAMA 3.3 with 70 billion parameters ^20^. The final stage transforms the PDF report into DICOM format for seamless integration with clinical imaging systems, converting it into an encapsulated DICOM object with preserved visual fidelity and appropriate DICOM metadata including study, series, and instance identifiers. DICOM-formatted reports integrate with hospital PACS, appearing alongside imaging studies in radiological workstations and benefiting from established healthcare data management protocols, ensuring longitudinal availability and security compliance.

### Cognitive Outcome Prediction

*Datasets*

Imaging and neuropsychological data from two non-overlapping datasets were used for outcome prediction analyses. The first dataset (training dataset) consisted of 604 patients from the Iowa Lesion Registry with lesion segmentations obtained via manual delineation on research-quality scans. The training set included 339 patients with ischemic stroke, 131 patients with hemorrhagic stroke, 75 patients with tumor resections, 30 patients with AVM resections, 11 patients with herpes simplex encephalitis, 9 patients with closed head trauma, 5 patients with penetrating head trauma, 2 patients with cyst resections, 1 patient with abscess resection, and 1 patient with limbic encephalitis. The second dataset (test dataset) consisted of 153 ischemic stroke patients with lesion segmentations obtained by applying the automated pipeline to clinical scans. Demographics for each dataset are shown in **Supplementary Table 6.**

**Supplementary Table 6. Demographics for outcome prediction datasets**

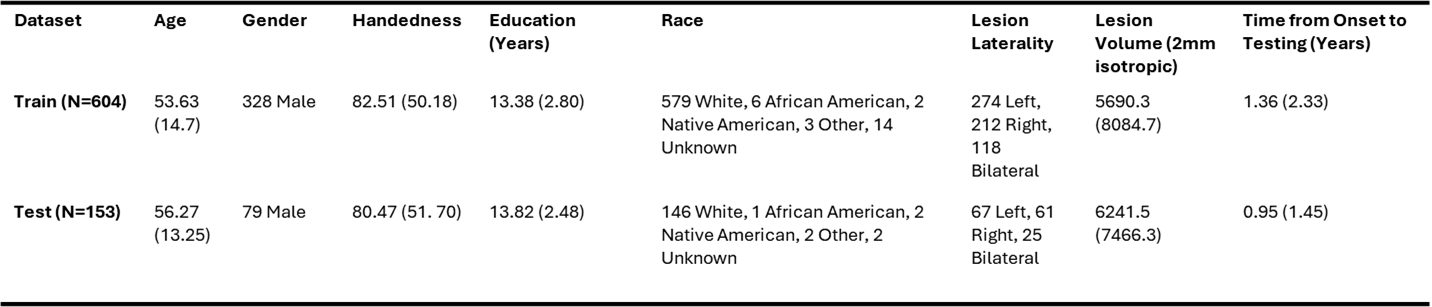

*Neuropsychological Outcome Measures*

Neuropsychological outcomes were selected on the basis of their being acquired in both the training and test datasets. This resulted in a set of 28 neuropsychological outcomes, consisting of the following (names followed by abbreviations used in main text):

1. WAIS Arithmetic (**arith**)
2. WAIS Similarities (**sim**)
3. WAIS Coding (**cd**)
4. WAIS Matrix Reasoning (**mr**)
5. WAIS Digit Span (**ds**)
6. WAIS Block Design (**bd**)
7. Chapman Reading Test (**chapman**)
8. Rey Complex Figure Test Recall (**cftr**)
9. Rey AVLT Trial 5 (**rey5**)
10. Rey AVLT Delayed Recall (**rey-dr**)
11. Rey AVLT Recognition Hits (**rey-hit**)
12. Trails A (**trails-a**)
13. Trails B (**trails-b**)
14. MAE Controlled Oral Word Association Test (**mae-cowa**)
15. MAE Sentence Repetition (**mae-sr**)
16. MAE Token Test (**mae-token**)
17. MAE Visual Naming (**mae-vn**)
18. MAE Aural (**mae-aural**)
19. MAE Reading (**mae-read**)
20. Boston Naming Test (**bnt**)
21. Benton Visual Retention Test (**brtc**)
22. Benton Test of Facial Recognition (**face-rec**)
23. Clinical Articulation Ratings (**clin-artic**)
24. Clinical Paraphasia Ratings (**clin-paraph**)
25. Clinical Fluency Ratings (**clin-fluen**)
26. Clinical Prosody Ratings (**clin-prosod**)
27. WRAT Reading (**wrat-read**)
28. Judgment of Line Orientation (**jlo**)

When possible, neuropsychological scores were adjusted for age or age and education using published norms and converted to population-normed *z*-scores ^21^. For each neuropsychological measure, patients were classified as either “impaired” or “not impaired”. For the MAE subtests, impairment thresholds were defined according to previously published cutoffs ^22^. For all other tests with population normed z-scores, the threshold for determining “impaired” status was defined as a population-normed z-score of -1.5 or below. Outcomes corresponding to clinician ratings of impairment, which are subjective clinician ratings determined according to clinical impression and include ratings of 1 (unimpaired), 2 (borderline), and 3 (impaired), scores less than 3 were considered “not impaired”, while scores of 3 were considered “impaired”. We note that per the Benton Clinical Handbook, ratings of 2 (borderline) are defined as: “...there is some question as to whether or not the performance is normal, but it is clearly not defective”. For the Benton Test of Facial Recognition, previously published cut-off scores (i.e. corrected score < 37) were used ^23^. The training and test sample sizes for each neuropsychological measure are shown along with impairment frequencies for each measure in **Supplementary Table 7**.

**Supplementary Table 7. Sample Sizes and Proportion Impaired**

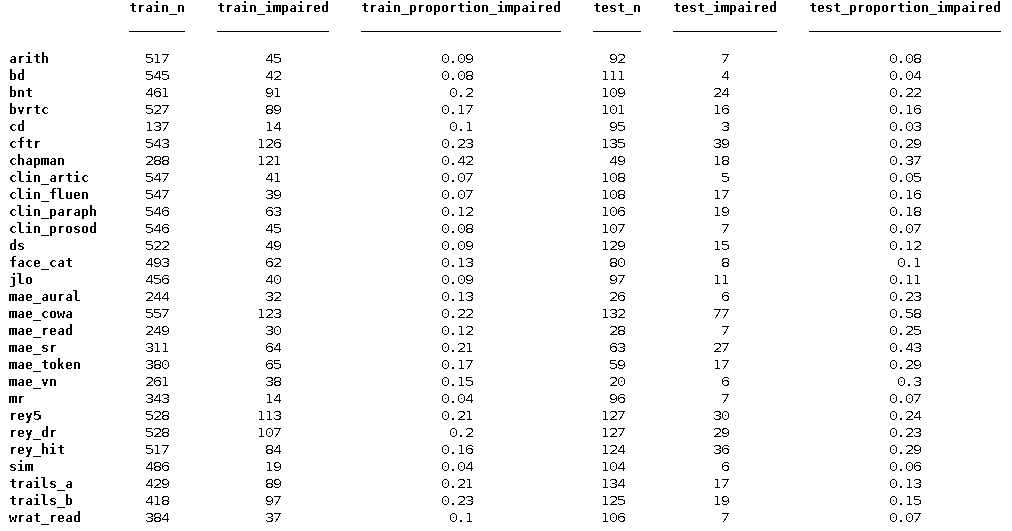

*Imaging Data*

For the training dataset, lesion masks were segmented and registered to the MNI-152 brain template as described previously ^24^. Lesion data were resampled to 2mm isotropic resolution. Structural lesion-network maps were generated with Lead-DBS by using each patient’s lesion as a seed region in deterministic tractography analyses of diffusion MRI data from the Human Connectome Project’s MGH 32-fold group connectome as reported in other publications ^25^, resulting in an estimated streamline disconnection map for each patient. Functional lesion-network maps were generated analogously by using each patient’s lesion as a seed region in resting-state functional connectivity analyses in the GSP-1000 normative subject sample described previously ^26^ using the principal component disconnection method ^27^, resulting in an estimated lesion functional connectivity map for each patient.

For the test dataset, lesion masks were automatically generated using the lesion prediction pipeline. Lesion-network maps were not generated for the test dataset as this functionality does not currently exist in the automated lesion prediction pipeline.

*Model Training*

Model training was performed using the Iowa Brain-Behavior Modeling Toolkit ^28^ in MATLAB r2022b (The MathWorks). We trained partial least squares (PLS) classification models using lesion segmentations and derived lesion-network features obtained from the training dataset and then applied these models to the test dataset to evaluate model performance. Hyperparameter optimization was performed using 5 repetitions of 5-fold cross-validation within the training dataset to determine the number of PLS components to include in each classification model using default settings in the Iowa Brain-Behavior Modeling Toolkit. Training and test folds for the hyper-parameter optimization were stratified by group (impaired vs. not impaired) to ensure representation of both groups in the training and test folds. As shown in **Supplementary Table 7**, there were large class imbalances for many of the neuropsychological outcome measures. To mitigate this during model training, we set the misclassification costs for patients with impairment proportional to the class imbalance (e.g., if there were twice as many patients without impairment, then the misclassification cost for patients with impairment was set to twice that of patients without impairment).

For each outcome, we generated one lesion location model, one structural lesion-network model, and one functional lesion-network model using the training dataset. The lesion location models were trained using voxel-based lesion masks as predictor features, while the structural and functional lesion-network models were trained using the derived structural and functional lesion-network maps. For models using structural and functional lesion-network maps, the input features were standardized to the range [0,1] and [-1,1], respectively, using centering and scaling factors defined in the training dataset. This resulted in four voxel-based PLS model coefficient maps for each outcome, one for the lesion location model, one for the structural lesion-network model, and one for the functional lesion-network model.

The automated lesion segmentation pipeline does not currently include the functionality required to generate lesion-derived network features, and so we could not directly apply the trained structural lesion-network and functional lesion-network models in the test dataset. To work around this limitation, we trained a second set of models, again in the training dataset, using lesion loads computed against the PLS model coefficient maps generated by the initial lesion-network models in the training dataset. Lesion loads on each lesion-network map were computed for each patient in the training dataset by taking the dot product between the patient lesion segmentations and the lesion-network model PLS coefficient map.

Finally, we trained “stacked” ridge-regularized logistic regression models that used the predicted class labels (impaired vs. unimpaired) from the lesion location model, the lesion loads on the structural lesion-network model coefficient map, and the lesion loads on the functional lesion-network model coefficient map as predictors. All features in the “stacked models” were standardized to range [0,1]. These “stacked” models therefore incorporated information about lesion location along with associated structural and functional networks. We note that while training secondary models using the lesion loads computed against the model coefficient maps generated from the training datasets is an inherently circular analysis within the training dataset, it should not bias evaluations of model performance in the fully independent test dataset since the test data were not used for generating or training either model. For estimates of cross-validation performance for the stacked models, lesion loads were computed using training fold coefficient maps (i.e. instead of the group-level coefficient maps) that were generated independently from the test folds, mirroring the approach used in the application to the independent test dataset and avoiding introducing biases into the cross-validation estimates.

*Model Evaluation*

Each trained single-modality model was then applied to the test dataset. For the lesion location models, the trained models were directly applied to the automatically generated lesion masks from the test dataset. For the models incorporating lesion loads on the lesion-network model PLS coefficient maps, input features for the test dataset were computed in the same way as for the training dataset (i.e., by taking the dot product between each test set patient’s vectorized lesion mask and the vectorized model coefficient map from the training dataset). For each neuropsychological test, this generated 4 predictions for each patient in the test dataset: one from the lesion location model, one from the structural lesion-network lesion load model, one from the functional lesion-network lesion load model, and one from the “stacked” model that combined the predicted class labels from the lesion location model with the lesion loads computed from the structural lesion-network and functional lesion-network model coefficient maps.

Model performance is reported in terms of the area under the ROC curve (AUC), which is robust even in the presence of large class imbalances such as those observed in the datasets under study, in line with recommendations for measurement of classification accuracy in human neuroimaging studies^29^. For descriptive purposes, we also report the classification accuracy for impaired patients and the classification accuracy for unimpaired patients using the classification score thresholds that optimized class separation in the training dataset. To evaluate the statistical significance of the observed classification accuracies at the trained thresholds, we performed permutation tests (1,000 permutation iterations) for each model and each outcome. These tests involved randomly shuffling the predicted group labels and then recomputing the AUCs to construct an empirical null distribution of AUC estimates. The observed classification accuracy was then compared against the empirical null distribution to obtain a p-value indicating the probability of obtaining an AUC at least as extreme as the observed value under the null hypothesis that the predicted classification scores contain no information about impairment status.

**Lesion Overlap Maps**

Lesion overlap maps for the training and test datasets are shown in **Supplementary Figure 4**.

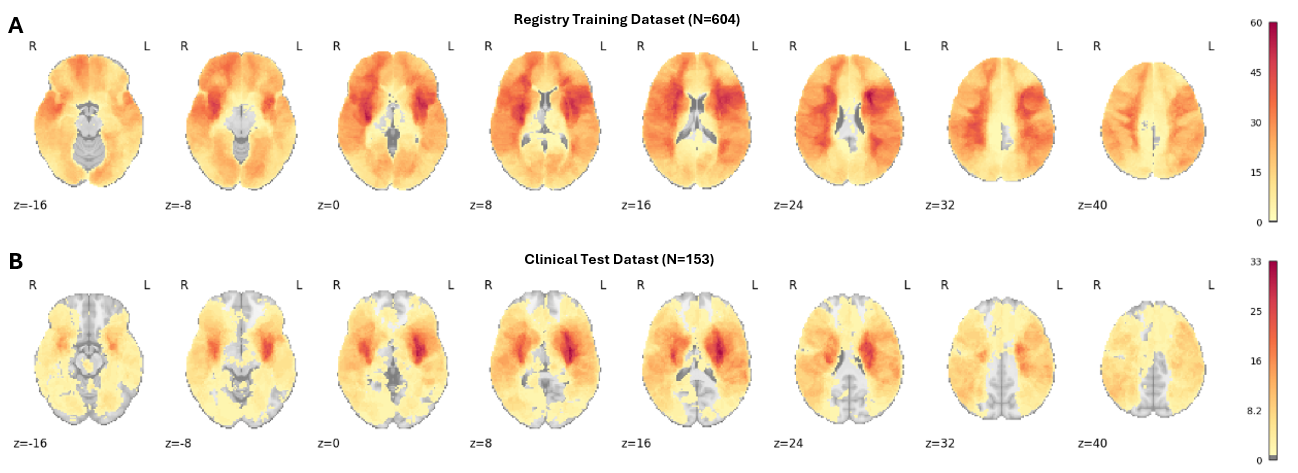

**Supplementary Figure 4.** Lesion Overlap Maps. **A**. Lesion overlaps for the training dataset. **B.** Lesion overlap maps for the test dataset. Voxel colors indicate the number of overlapping lesions at each voxel.

**Cross-Validation Results**

The results of cross-validation analyses within the training dataset are shown in **Supplementary Figure 5**.

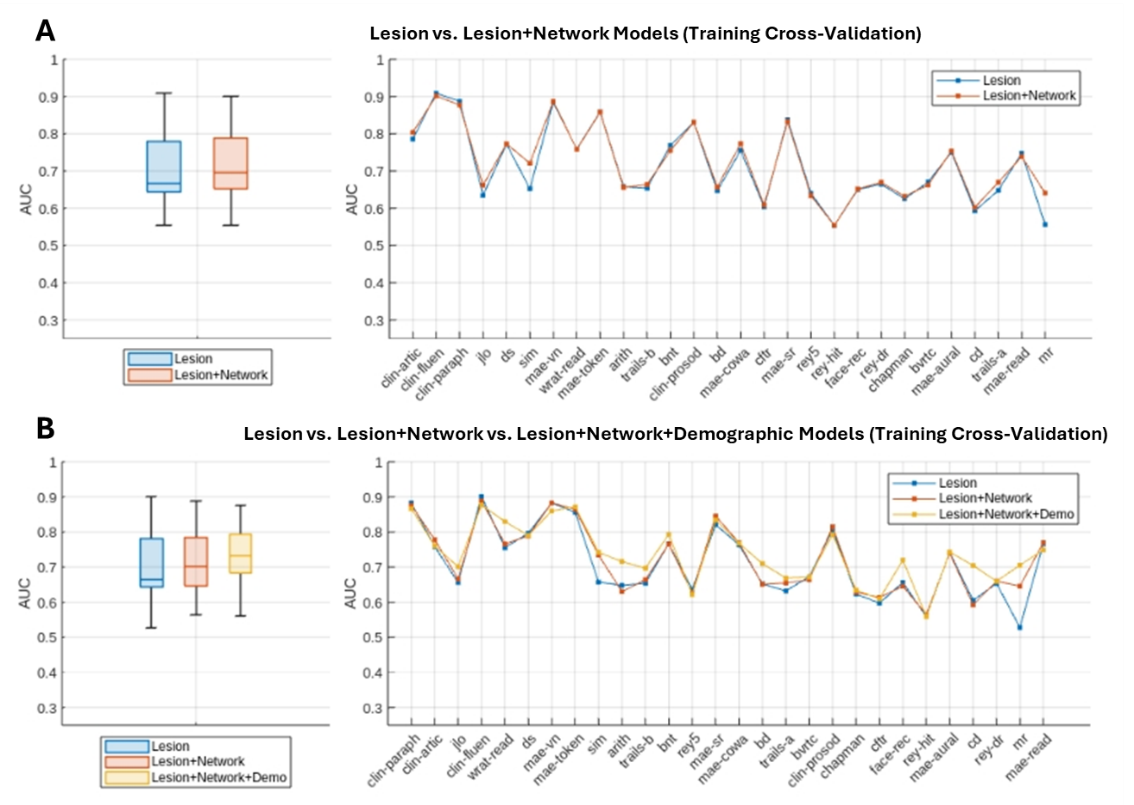

**Supplementary Figure 5.** Cross-validation performance in the training dataset. **A.** Cross-validation performance for 5 repetitions of 5-fold cross-validation is shown for the lesion models and lesion + network models in the training dataset. AUC values correspond to the average cross-validation test set AUC across folds and repeats of the cross-validation procedure. Cognitive outcomes in the right panel are sorted identically to **Figure 4A**, highlighting differences in relative performance in training dataset cross-validation vs. out-of-sample prediction in the independent test dataset. The rank correlation between the lesion + network model AUCs obtained from training dataset cross-validation and the lesion + network AUCs obtained from the independent test dataset was moderate (r=0.56). **B.** Same as (A), but with the addition of lesion + network + demographic models. The rank correlation between the lesion + network model AUCs obtained from training dataset cross-validation and the lesion + network AUCs obtained from the independent test dataset was moderate (r=0.53). Cognitive outcomes are sorted identically to **Figure 4C**.

**Concordance of Predictions between Manual and Automated Lesion Segmentations**

The concordance of predictions between manual and automated lesion segmentations is shown in **Supplementary Figure 6**.

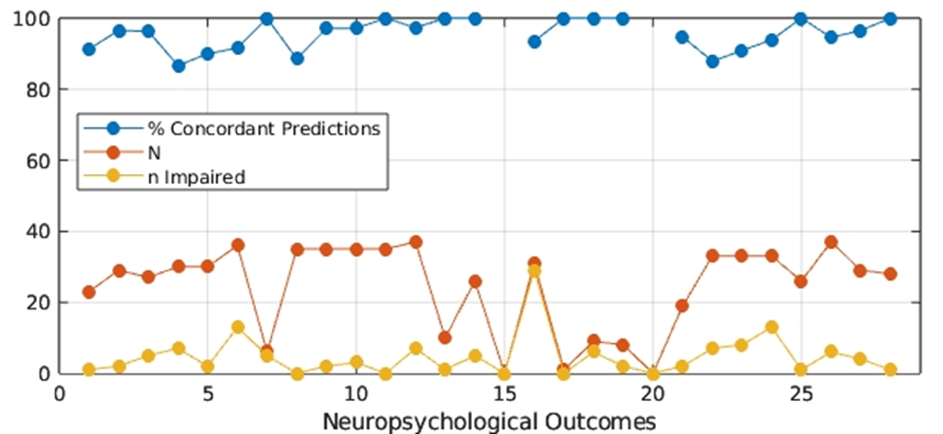

**Supplementary Figure 6.** Concordance of predictions for test dataset patients with manual lesion segmentations. For test dataset patients that also had manually segmented lesion images (N=57, 681 total predictions), the trained lesion-only models were applied to both the manual lesion segmentations and the automated pipeline lesion segmentations to obtain two sets of predicted class labels – one for the manual lesion segmentations and one for the automated lesion segmentations. The concordance between the predicted class labels obtained from the automated vs. manual lesion segmentations was then measured for each neuropsychological outcome and across all outcomes simultaneously. The percentage of predictions that were concordant between the manual and automated lesion segmentations is shown for each outcome (blue line) along with the sample size of patients with ground truth lesions (orange) and the number of patients with ground truth lesions with impairment for each outcome (yellow). Across all outcomes, the average concordance rate was 96% (SD=4%), with a range of 87%-100%. Across all aggregated predictions (i.e. across all outcomes; 549 total predictions) for unimpaired patients, the concordance rate was 95%. Across all aggregated predictions (i.e. across all outcomes; 132 total predictions) for impaired patients, the concordance rate was 94%. These results show that for the 57 test dataset patients with ground truth manual lesion segmentations, the predicted class labels were highly consistent regardless of whether the ground truth manual lesion segmentations vs. automated pipeline lesion segmentations were used to obtain the predictions.

**System Evaluation**

**Design and Implementation.** The system’s architecture was designed to support seamless integration into clinical workflows, ensuring compatibility with hospital Picture Archiving and Communication Systems (PACS) through standard DICOM protocols. Reports are generated as DICOM-encapsulated PDF files, enabling direct incorporation into patients’ imaging records and accessibility via radiological workstations, thus embedding stroke analysis results within existing imaging workflows and minimizing the need for additional systems or manual processes. The deployment strategy leverages Docker containerization, splitting the core processing pipeline and language model services into separate containers for enhanced flexibility, security, and scalability. The large language model, hosted locally in a dedicated container using the Ollama framework, processes only extracted features from the pipeline, ensuring that sensitive patient data remains confined to the primary processing container and is never exposed to external services. This design supports compatibility with HIPAA requirements by preventing data leakage and enabling secure, on-premises deployment, while the containerized architecture dynamically adapts to available computational capabilities, supporting both GPU-accelerated and CPU-based processing to meet diverse clinical environment needs. Validation testing in a simulated hospital PACS environment confirmed the system’s ability to handle automated processing triggers, real-time status monitoring, and secure data purging post-analysis, demonstrating its readiness for enterprise-scale clinical integration. A custom profiler class monitors execution time, memory, and resource usage, logging every run for traceability, while comprehensive logging records execution flow, timing, and exceptions without including sensitive patient information. The codebase achieved 92% source code coverage through unit tests, complemented by integration tests to prevent unexpected errors, ensuring reliability for clinical deployment. The modular design separates concerns across pipeline stages—DICOM processing, lesion segmentation, outcome prediction, and report generation—using standardized data formats for communication, enabling robust quality assurance, preventing error propagation, and allowing independent testing and validation of each module. The system adheres to Brain Imaging Data Structure (BIDS) for data organization, ensuring reproducibility and compatibility with neuroimaging tools, and leverages established frameworks, including ITK for medical image processing, PyDICOM for DICOM operations, and PyTorch with MONAI for deep learning components, benefiting from community support and standardized interfaces. All deep learning models and processing pipelines use fixed random seeds for deterministic, reproducible results, except for the registration module, where minor variations from iterative optimization have negligible downstream impact. Development followed Agile methodology, using GitHub for collaboration, with strict code review and automated validation via GitHub Actions, while documentation, hosted on a GitHub Wiki, includes API references, system architecture, testing protocols, and deployment guides, ensuring clarity for development and deployment teams.

**Runtime Analysis.** We report results from a single, fully automated processing run on an out-of-sample clinical test dataset comprising 160 stroke patients. All cases met minimum eligibility criteria (one structural image, DWI, and ADC), and the system successfully processed all subjects end-to-end without triggering any fail-safe mechanisms. All intermediate outputs (image classification, brain masks, registrations, lesion segmentations) were visually reviewed for quality assessment.

Performance benchmarks were conducted on a workstation with an Intel Xeon w7-3545 CPU (24 cores, 48 threads, 4.8GHz max frequency). Most components ran on a single CPU core, except registration, which utilized 24 cores due to its computational intensity. As shown in **Table 1**, CPU-only processing achieved an average end-to-end time of 274.8 seconds (approximately 4.6 minutes), ranging from 188.1 to 472.5 seconds (SD=47.0). Registration was the most time-intensive step at 85.7 seconds on average, followed by report generation at 146.3 seconds, reflecting the sequential nature of the pipeline on the CPU. Enabling GPU acceleration on an NVIDIA RTX 6000 Ada (48GB memory) reduced the average end-to-end processing time by 55.9% to 121.3 seconds (approximately 2 minutes), with 95% of cases completing in under 3 minutes (range=80.4 to 238.1 seconds, SD=22.3). Registration time decreased by 10.5% to 76.7 seconds, while report generation saw the most significant improvement, dropping 93% to 10.0 seconds, mainly due to GPU acceleration of large language model inference for text generation. This efficiency, detailed in **Table 1**, underscores the GPU’s value for rapid analysis in acute stroke care settings.

**Supplementary Table 8.** Runtime summary of processing system components in seconds.

| **System Component** | **CPU** | | | | **GPU** | | | |
| --- | --- | --- | --- | --- | --- | --- | --- | --- |
|  | **Mean** | **Min** | **Max** | **Std** | **Mean** | **Min** | **Max** | **Std** |
| Data Selection and Preprocessing | 36.1 | 26.9 | 68.0 | 7.0 | 29.6 | 18.8 | 48.0 | 5.0 |
| Registration | 85.7 | 53.8 | 197.4 | 17.4 | 76.7 | 49.1 | 168.8 | 15.4 |
| Lesion Segmentation | 5.7 | 4.9 | 10.8 | 1.0 | 4.1 | 3.9 | 4.6 | 0.2 |
| Outcome Prediction | 1.0 | 0.9 | 1.1 | 0.0 | 0.9 | 0.8 | 0.9 | 0.0 |
| Report Generation | 146.3 | 101.6 | 195.2 | 21.6 | 10.0 | 7.8 | 15.8 | 1.65 |
| **Total** | **274.8** | **188.1** | **472.5** | **47.0** | **121.3** | **80.4** | **238.1** | **22.3** |
